# Geography, Ancestry, Age and Sex Shape Somatic Autosomal Mosaic Chromosomal Alterations in Blood

**DOI:** 10.1101/2025.10.19.25338239

**Authors:** Jasmine Ryu Won Kang, Yeekyung June Kim, Kimberly Skead, David Soave, Jonathan Evans, Vanessa Bruat, Michelle P. Harwood, Quaid Morris, Enock Matovu, Julius Mulindwa, Harry Noyes, Annette McLeod, Scott Hazelhurst, Zane Lombard, Michele Ramsay, Marie-Julie Fave, Philip Awadalla

## Abstract

Clonal hematopoiesis, through the age-associated accumulation of somatic mutations in blood, is strongly associated with hematological malignancies and other chronic diseases. These mutations have largely been characterized in individuals of European ancestry or environments, and consequently, it remains unclear how mutation-recurrence patterns vary across populations of different histories or non-European ancestries. Here, we evaluate how variation in geography, ancestry, genetics and sex shape the prevalence of mosaic chromosomal alterations (mCAs) among 47,369 individuals from the Canadian Partnership for Tomorrow’s Health, including a founding population cohort in Quebec, and 13,562 individuals from within South, Central, West and East Africa though the H3Africa consortium. We identified autosomal mCA hotspots that were ancestry- and sex-specific, mapped novel ancestry-specific germline variants associated with autosomal mCA prevalence, and estimated heritability rates to quantify the germline genetic contribution to autosomal mCA variance. We also showed how mCAs impact blood transcriptomes, implicating stabilizing selection as a mechanism by which copy number gain mutations are tolerated in healthy blood. Collectively, mapping the landscape of autosomal mCAs in populations of diverse ancestry illustrates similarities but also highlights important ancestry, geographic, and sex-specific differences.

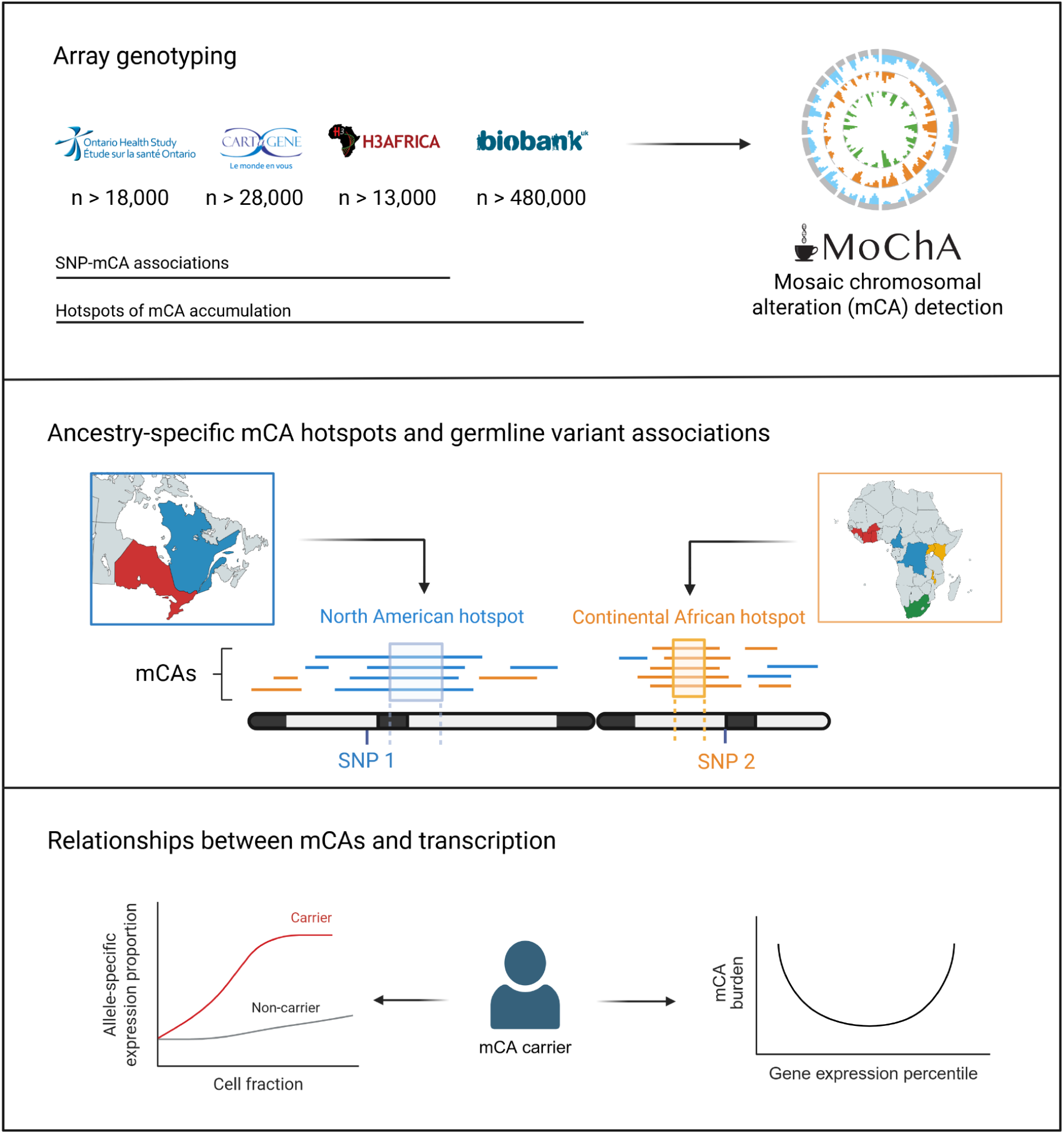

## Introduction

Clonal hematopoiesis (CH) arises when a hematopoietic stem cell (HSC) acquires a somatic mutation that provides that clone a fitness advantage over other clones, causing an expansion of the mutant clone to detectably elevated frequencies in the blood^1–3^. CH can be observed by assaying somatic variation of point mutations at candidate driver genes, such as *DNMT3A* and *TET2*^4^, or of large structural mutations, known as mosaic chromosomal alterations (mCAs)^5^. Both point mutation and mCA-driven CH are considered critical pre-disease states or risks for a wide range of pathologies^6–10^, most notably in blood malignancies^4,9,11–15^ and cardiovascular diseases^16,17^, although most CH drivers do not result in adverse outcomes. CH is also a ubiquitous feature of the aging HSC compartment, and has until recently been predominantly characterized in populations of European ancestry^11,18,19^. The landscape of mCAs, including their preferential accumulation in genomic hotspots and potential germline determinants, remains largely underexplored among individuals of diverse ancestral or genomic backgrounds, such as continental African populations or ‘founding’ populations.

Previously, whole-genome sequencing in 67,390 individuals in the National Heart, Lung, and Blood Institute Trans-Omics for Precision Medicine (TOPMed) revealed that individuals of European ancestry had the highest rates of autosomal mCAs, compared to individuals of African-American ancestry^20^. While the TOPMed cohort includes African-American individuals living in the United States, the H3Africa consortium^21^ includes individuals of African ancestry living throughout Africa, of different ethnicities and cultural or language groups. Given the high degree of genetic and environmental diversity within continental Africa^22,23^, there may exist regional differences in somatic mutation acquisition, with specific germline factors potentially associated with these differences.

The Canadian Partnership for Tomorrow’s Health (CanPATH)^24^ also enables a view of mutation recurrence patterns driven by regional subpopulation structure, particularly as we leverage genotyping data from individuals of European ancestry living in Ontario, but also including participants from Quebec, which is a founder population including individuals of French ancestry colonizing ‘New France’, with a unique genetic architecture characteristic of founding events and population growth^25–28^.

Here, we present a genomic characterization of 1,558 autosomal mCAs from a total of 56,940 participants of diverse ancestries, including continental African populations and a founder population of European ancestry. We determined overall mCA prevalence rates comparing ancestries and identified mCA hotspots that differ by both ancestry and sex. Our approach to locate mCA hotspots provides a scalable framework to compare autosome-wide mCA accumulation patterns across ancestries, while accounting for chromosome-specific mCA propensities. While prior studies have identified germline variants associated with mCA risk^12,20,29^, we discovered novel variants specifically associated with hotspot-overlapping mCAs occurring in *cis*, as well as those associated with autosome-wide mCAs. Specifically, we identified unique genetic associations within populations of different ancestries, including among sub-populations in continental Africa, and also among subpopulations of European ancestry, including in Quebec. Finally, in a subset of European-ancestry participants, we query the impact of mCAs on bulk transcriptomes in blood, we show how such large-scale somatic mCAs are impacting molecular phenotypes, and offering hypotheses why these mCAs may be tolerated.

## Results

### Autosomal mCAs were profiled in 56,940 individuals across continental African populations and European ancestry populations

To profile patterns of mCA accumulation across different ancestries, we leveraged population cohort data from Ontario (Ontario Health Study^30^, OHS), Quebec (CARTaGENE^31^, CaG), continental Africa (AWI-Gen^32,33^ and TrypanoGEN+^34^), and the UK (UK Biobank^35^, UKB), genotyped with the Axiom, GSA, H3Africa, and Axiom/BiLEVE arrays, respectively (Figure 1). The AWI-Gen and TrypanoGEN+ cohorts include ten countries spanning West, East, Central, and South Africa. We identified mCAs using MoChA^11^, a well-established approach that has been applied in numerous population cohorts^12,13,20,36,37^, and determined the prevalence of mCAs across our cohorts. To ensure valid comparisons across cohorts, regions, or ancestries, and to minimize biases in mCA detection due to variable genotyping array densities, we used genotyping datasets in which SNPs were randomly “downsampled” to match the SNP density of the arrays with the lowest densities (∼600,000 SNPs), namely the GSA and Axiom arrays. We noted that arrays with a higher SNP density (H3Africa) detect a lower median mCA length compared to arrays with lower SNP density (GSA and Axiom) (p < 2.2e-16, Kruskal-Wallis test), presumably driven by the greater SNPs density with larger SNOP content in H3Africa arrays, which allows for the detection of mCAs of smaller length. To assess how array types may impact mCA calls, we also compared mCAs called using the 925 samples within the Quebec cohort which had been genotyped with both the GSA and the OMNI Illumina arrays. Of the 27 samples carrying autosomal mCAs in CaG-GSA among the GSA/OMNI overlapping samples, 23 (85%) were also found to carry mCAs using the OMNI array (Table S1). The cell fractions of these mCAs demonstrated correlation between GSA and OMNI (R^2^ = 0.83, linear regression). Finally, given previous reports that X chromosome mCAs differ significantly in overall prevalence and copy change types across technologies^20^, we chose to focus our efforts in the current analyses on mCAs occurring on autosomes. To allow a direct comparison of mCA occurrence across ancestries, we only retained samples of European ancestry in the North American cohorts, and all samples in the continental African cohorts were of non-admixed ancestry (Figure S1). The overall prevalence of autosomal mCAs in each of the cohorts were as follows: North America: 2.7% in OHS, 2.5% in CaG-GSA; continental Africa: 2.5% in AWI-Gen, 2.0% in TrypanoGEN+; Europe: 3.5% in UKB (Figure 1).

**Fig. 1.**
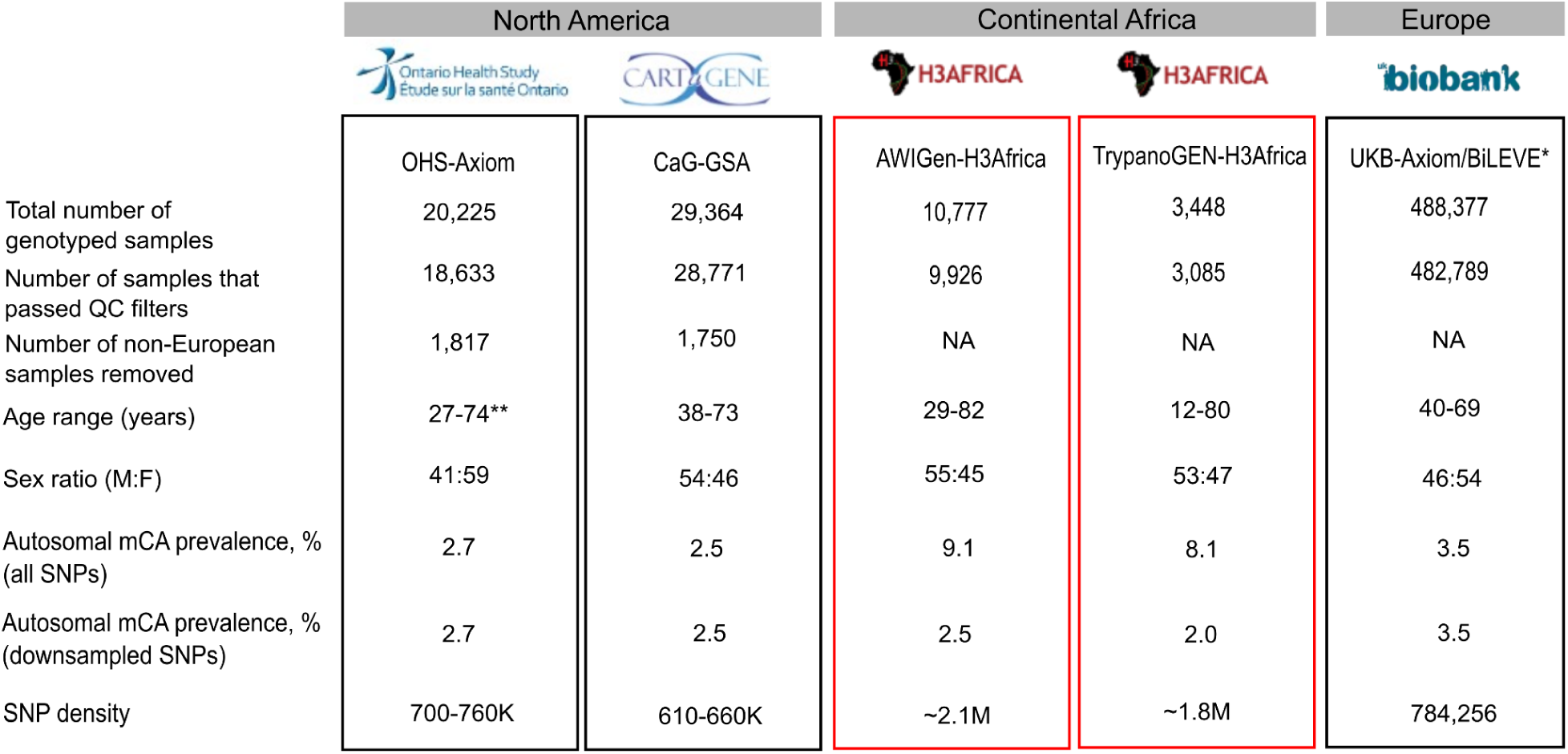
Sample counts, demographics, and mCA prevalence in the North American, continental African, and European population cohorts. mCA prevalence is defined as the percent of samples with at least one autosomal mCA, and was calculated either from mCAs called with all SNPs in a given array or with a set of downsampled SNPs. Boxes outlined in red represent cohort-arrays that were downsampled in SNP density to match that of the lowest-density arrays (GSA and Axiom, ∼600,000 SNPs). All samples of non-European ancestry were removed from OHS and CaG. OHS=Ontario Health Study, CaG=CARTaGENE, UKB=UK Biobank, GSA=Global Screening Array. *All metrics for UK Biobank are reported in Loh et al.^12^ **Age data is available for the first sequencing batch of OHS.

### Age-associated changes in autosomal mCA prevalence are common across ancestries, while chromosome-level mCA prevalence is different across ancestries

We conducted an age-stratified analysis of mCA prevalence and cell fraction (including all SNPs in a given array) for all participants in AWI-Gen and the North American cohorts. The median age ranged from 51 years in AWI-Gen to 55 years in North America, and among samples carrying mCAs, most samples carried only one mCA. Consistent with previous literature, we found that mCA prevalence increased with age (p < 2.2e-16, two-sample proportion test, <=40 vs >70 years) (Figure S2A). However, we also found that mCA cell fraction decreased with age, whereby the cell fractions in mCAs detected in individuals over 70 years old were significantly lower than those in the 61-70 year group (p = 0.034, Wilcoxon rank-sum test; Figure S2B). We analyzed North American cohorts and AWI-Gen sub-populations separately and similarly found a significant decrease in cell fraction with age in AWI-Gen (p=0.037, Kruskal-Wallis test) (Figure S2C-F). We stratified this analysis by sex and mCA type, and found similar increases in mCA prevalence with age in both sexes and in all mCA types; however, the decrease of mCA cell fraction with age was only significant in males (p = 2.5e-3, Kruskal-Wallis test) and in CN-LOH events (p = 0.022, Kruskal-Wallis test). These results may suggest the effect of a small number of HSCs dominating hematopoiesis in older age, consequently compressing the cell fractions of the remaining clones.

We next compared mCA prevalence across chromosomes using mCAs called from the downsampled SNPs to account for disparities between array probe densities. Averaged across all cohorts, chr 8, 18, and 21 were least frequently impacted by mCA events (1.7%, 2.1%, and 1.5% of total autosomal mCAs, respectively), while chr 1, 11, and 13 were most frequently impacted (6.6%, 7.1%, 6.2% of total autosomal mCAs, respectively). We visualized the genomic locations of mCA occurrence, normalized by cohort size and stratified by mCA type (Figure 2). For example, we observed a high normalized frequency of gain mCAs on chr 5 in the continental African cohorts, which was not observed in the North American cohorts, nor in the UKB or TOPMed cohorts, the latter of which includes individuals of African-American individuals^20^, suggesting that this chr 5 mCA accumulation is specific to participants recruited from continental African cohorts. In contrast, chr 13 was frequently impacted by CN-LOH mCAs in the two North American cohorts and the UKB cohort, while it was less frequently impacted by these events across the continental African cohorts.

**Fig. 2.**
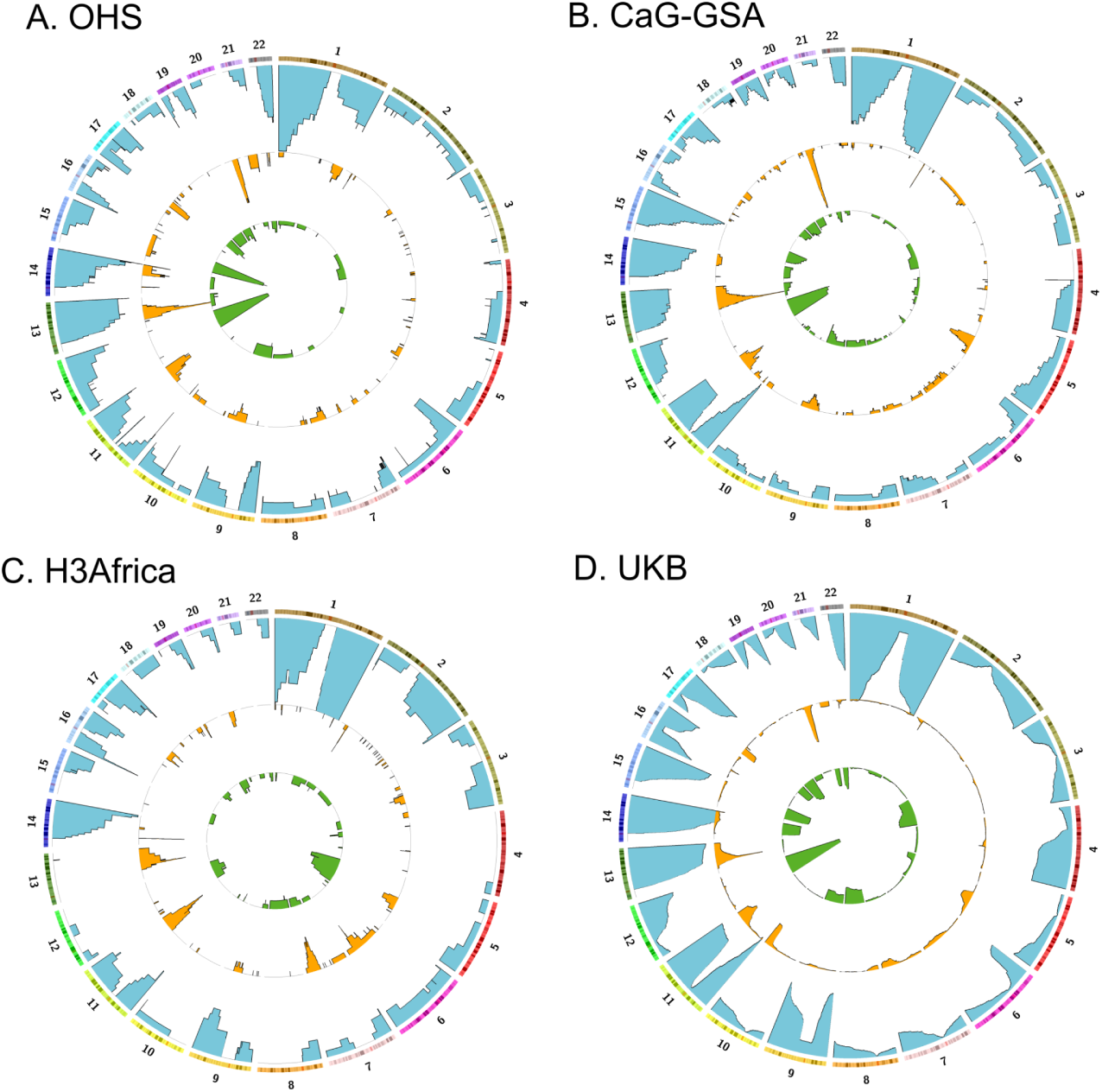
Genomic distributions of autosomal mCAs. mCAs were profiled in OHS, n=517 (A), CaG-GSA, n=726 (B), H3Africa, n=351 (C), and UKB, n=14,292 (D). mCAs were called using SNPs downsampled to the least-dense array. mCAs are divided by copy change type in each track (Gain=green, Loss=orange, CN-LOH=blue). Chromosomes are labeled on the edge of the Circos plot. Centromeric regions are represented in red. The height of the bars represents the proportion of all mCAs detected in each cohort. H3Africa combines all African participants from different geographic regions. All samples of non-European ancestry were removed from OHS and CaG.

### Autosomal mCA hotspots vary across populations and continents

Given that certain chromosomes have higher rates of autosomal mCA prevalence than others, we next statistically characterized autosome-wide hotspots of mCA accumulation while accounting for potential confounding effects of chromosome-specific sensitivity for mCA events. Different chromosomes have different propensities for somatic structural variation, owing to factors such as chromosome size, chromatin structure, and gene content^38^. To compare mCA accumulation across ancestries, we employed right-tailed binomial tests to identify genomic positions that are significantly more frequently impacted by mCAs than chromosomal baseline levels, again using downsampled SNPs.

We observed 13 significant hotspot regions in Ontario, 16 in Quebec, 29 in UKB, and 10 in the continental African cohorts, where every queried SNP within a hotspot region had a significant binomial test and mCAs were called with downsampled SNPs (Figure S3). For each hotspot region, we queried whether any part of that region was found as a hotspot in other cohorts or populations. We note that hotspot identification is sensitive to cohort size, as smaller cohorts may have less stable estimates of baseline mCA prevalence. Thus, we assess hotspot region overlap as a binary variable, rather than quantify the degree of overlap, for the purpose of locating cohort-specific hotspots (ie. hotspots that did not overlap to any degree with a hotspot in any other cohort). We identified two unique hotspot regions, one on chr 10 that was exclusively found in Ontario and the other on chr 4 among individuals in the founding population in Quebec (Figure S3A-B). We also identified three hotspot regions unique to the continental African cohorts on chr 4, 6, and 8 (Figure S3C).

We found that mCAs specifically overlapping hotspot locations had a greater bias towards CN-LOH and loss events, rather than gain events, in our cohorts as has been previously observed^11–13,20^. However, mCAs overlapping hotspot locations also had a significantly lower median cell fraction compared to mCAs overall (0.10 in mCAs overlapping hotspots, 0.12 in overall mCAs, p = 3.6e-04, Wilcoxon rank-sum test). Overall, these results suggest that mCAs overlapping hotspots have distinct characteristics that distinguish them from the general mCA pool.

Given the high degree of genetic diversity within the African continent^39,40^, we asked whether geographic region-specific differences in mCA hotspots could be detected among the continental African cohorts. We subdivided the African cohorts into four geographic regions: West, East, Central, and South^21,32^ (Table S5). For each hotspot region across all autosomes, we noted the mCAs overlapping the hotspot and the geographic region from which the sample containing the mCA was derived. Initially, we attempted to identify hotspots independently within each African geographic region; however, sample size limitations precluded such analyses. Instead, we categorized the hotspot based on the number of geographic regions represented by the mCAs overlapping the hotspot and found that no hotspot was exclusively derived from one geographic region. Rather, most hotspots were overlapped by mCAs from three or four geographic regions in continental Africa. Using downsampled SNPs, we are mitigating against differences in platform content and technology, but we may also be biased towards finding shared hotspots.

When using all assayed SNPs to discover potential novel hotspots, we found 10 out of 33 hotspot regions in continental Africa that were not found in the UKB or North American cohorts (Table S2).

We observed three hotspot regions that were consistently shared across all cohorts (regions on 11q, 13q, and 16p) (Figure S3). Notably, the 13q hotspot region encompasses *DLEU2*, which encodes a long-noncoding RNA implicated in tumorigenesis, affecting cell proliferation, migration, invasion, and apoptosis^41^, and is a well-characterized driver mutation in chronic lymphocytic leukemia (CLL), along with 13q CN-LOH and trisomy 12^42,43^. Both 13q loss and CLL are less common in individuals of East Asian ancestry compared to individuals of European ancestry^13,44^. In our dataset, we also observe that 13q loss events occur at approximately half the frequency in individuals of African ancestry compared to those of European ancestry (p = 4.50e-5, two-sided χ^2^). These data suggest that the acquisition of mCAs overlapping 13q is more common among populations of European ancestry, consistent with the observation that CLL is rarer among African ancestry populations compared to those of European ancestry^45,46^.

### Autosomal mCA hotspots localize near CH driver genes and demonstrate sex specificity

Hotspots of mCAs were identified in proximity to several loci previously associated with CH, or known CH drivers, including *DNMT3A, TET2,* and *ASXL1*^47^ (Figure 3 and Table S3). The p-values associated with these specific hotspots surpassed multiple testing correction among the North American cohorts, but not so in the continental African populations. Additional hotspots also localized near the *IGH* locus, which undergoes somatic VDJ recombination for immunoglobulin production^48^. We also observed mCA hotspots overlapping *ABCA1*, a gene in which functionally deleterious alleles have been previously implicated in high-density lipoprotein (HDL) deficiency^49^. In both European- and African-ancestry cohorts, we identified significant mCA hotspots overlapping SNPs in the 16p region which contains a cluster of hemoglobin subunit loci, including *HBZ, HBA1, HBA2, HBM,* and *HBQ1*, which play essential roles in oxygen transport, potentially implicating this mCA with either clonal or individual fitness^50^.

**Fig. 3.**
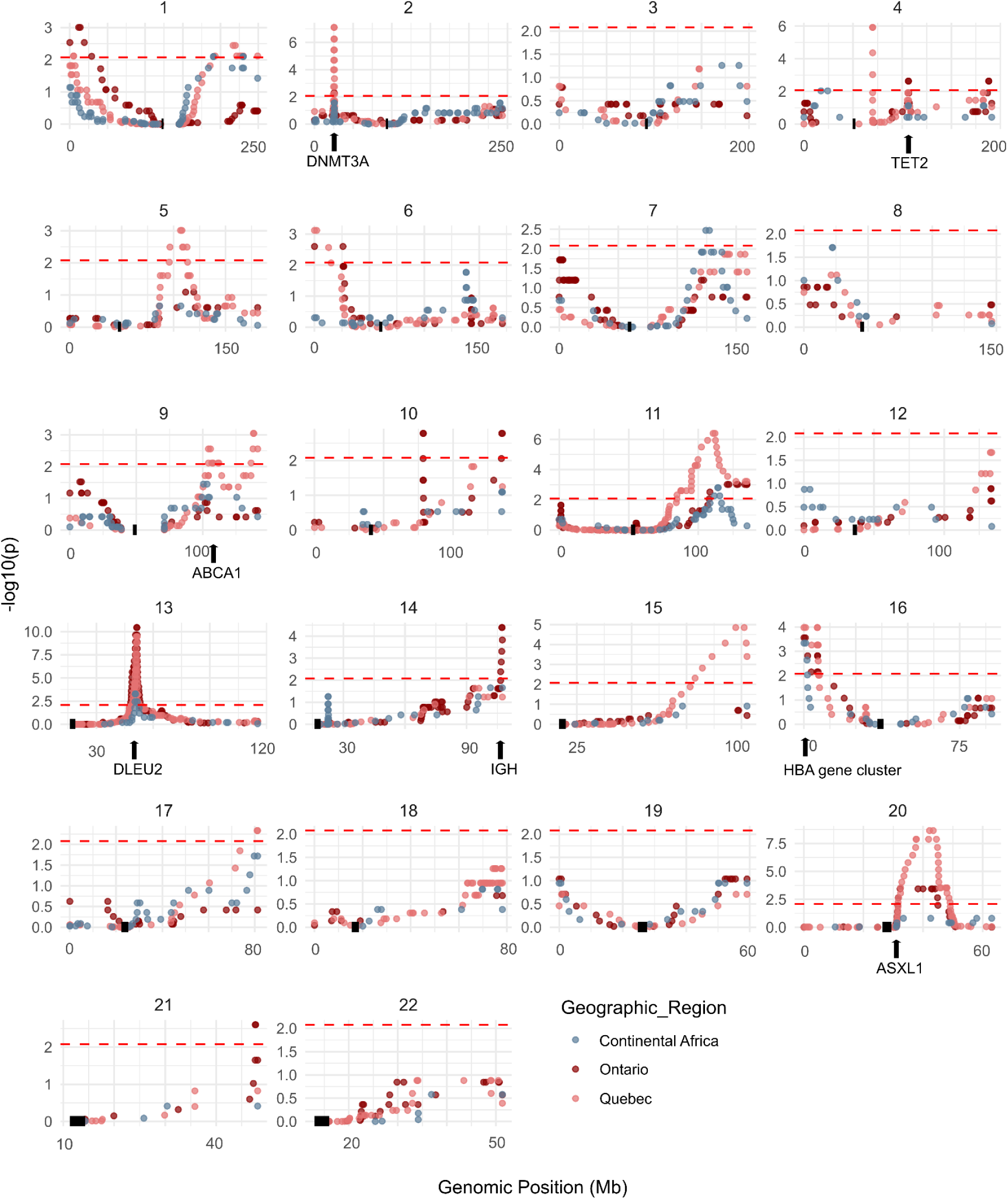
mCA hotspots localize near the Hb A gene cluster, *ABCA1*, *IGH*, and CH driver genes. mCA calls per chromosome were derived using SNPs downsampled to match the least dense arrays. Genes of interest nearby hotspot regions are indicated along the position axis. For some peaks, there were no known key genes of interest surrounding the hotspot. Centromeric regions in each chromosome are indicated in solid black. The significance threshold for hotspot identification after Benjamini-Hochberg multiple testing correction is shown as a red dotted line (adjusted α = 0.0084), adjusting for all binomial tests in Ontario, Quebec, and continental Africa. All samples of non-European ancestry were removed from Ontario and Quebec. See also Fig. S3-4 and Tables S3.

The mCA prevalence rate has been previously reported to differ across biological sex for both autosomes^20^ and X chromosomes^51^. In our data among the North American cohorts, we observed female-specific hotspot regions on chr 1, 4, 6, 9, 10, 11, 14, 15, 16, and 21, as well as male-specific hotspot regions on chr 9, 11, and 16 in Quebec (Figure S5A-B). We also identified sex-specific hotspots in continental Africa, some of which were shared with sex-specific hotspots in North America, while others were unique to Africa. We observed male-specific hotspots on chr 9 and 11, which overlapped with male-specific hotspots in Quebec (Figure S5D,F). In addition, we observed female-specific hotspots on chr 1 and 14, which overlapped with female-specific hotspots in Quebec and Ontario, respectively (Figure S5A,C,E). Notably, the chr16 hotspot was seen to be female-specific in Ontario, Quebec, and Africa, and overlaps the HbA gene cluster, suggesting sex-specific fitness advantages of somatic structural variation within these loci.

### Novel cis and autosome-wide germline variants in North America and West Africa are associated with mCAs

We next aimed to understand whether mCAs, either accumulating in hotspots or those found autosome-wide, in our separate population cohorts are associated with genetic variation. Several studies have described germline variants in association with mCA acquisition and clonal expansion^11–13,20^. Here, we tested for associations between germline variants and mCA presence, independently in each sub-population or cohort, and compared the significant associations across ancestries or to previous studies. We performed Fisher’s Exact Test (FET) to discover associations between specific germline genotypes and the following four phenotypes: loss, gain, CN-LOH and type-agnostic events (any event type at a given location), and performed replication analyses within each population. *Cis* association tests were performed following the methodology of Loh et al.^11^ and leveraging hotspots identified in the previous analysis, defining cases as the presence of a hotspot-overlapping mCA within a 4Mb window of a variant within a cohort. As mCA hotspots called from downsampled SNPs could be biased to identifying hotspots shared between ancestries, all association analyses were performed using hotspots and mCAs called from all SNPs. These analyses were performed on each of the North American cohorts and the West, East, and South African cohorts, while the Central African dataset was excluded from any association analyses due to insufficient sample size.

We were able to identify and replicate unique *cis* associations of germline variants with mCA presence within the Ontario and Quebec cohorts (OHS and CaG-GSA) and the West African cohort (Figure 4A-C, Table 1). All significant and replicated associations have not been previously reported. In OHS, intronic variants in the *LRMDA* locus and variants in the 3’ UTR region of the *KCNMA1* locus were identified as being associated with CN-LOH and type-agnostic events overlapping a hotspot. *KCNMA1* has been characterized as a tumor suppressor gene in gastric cancer^52^ and amplification of the locus drives cell proliferation in prostate cancer^53^. In the Quebec cohort, a non-coding variant was associated with type-agnostic events (rs72759877, p < 1.62e-12), and an intronic variant mapping to a long non-coding RNA (rs72759899, p < 1.61e-9), *LINC02348*, was associated with CN-LOH events and type-agnostic events. In the West African cohort, *cis*-variants associated with mCA events of any type were identified on chr 14 in intronic regions of the *OR4K1* gene. These variants were found to segregate in other African cohorts albeit at reduced frequencies (MAF=0.02-0.06) than observed in West Africa (MAF=0.06).

**Fig. 4.**
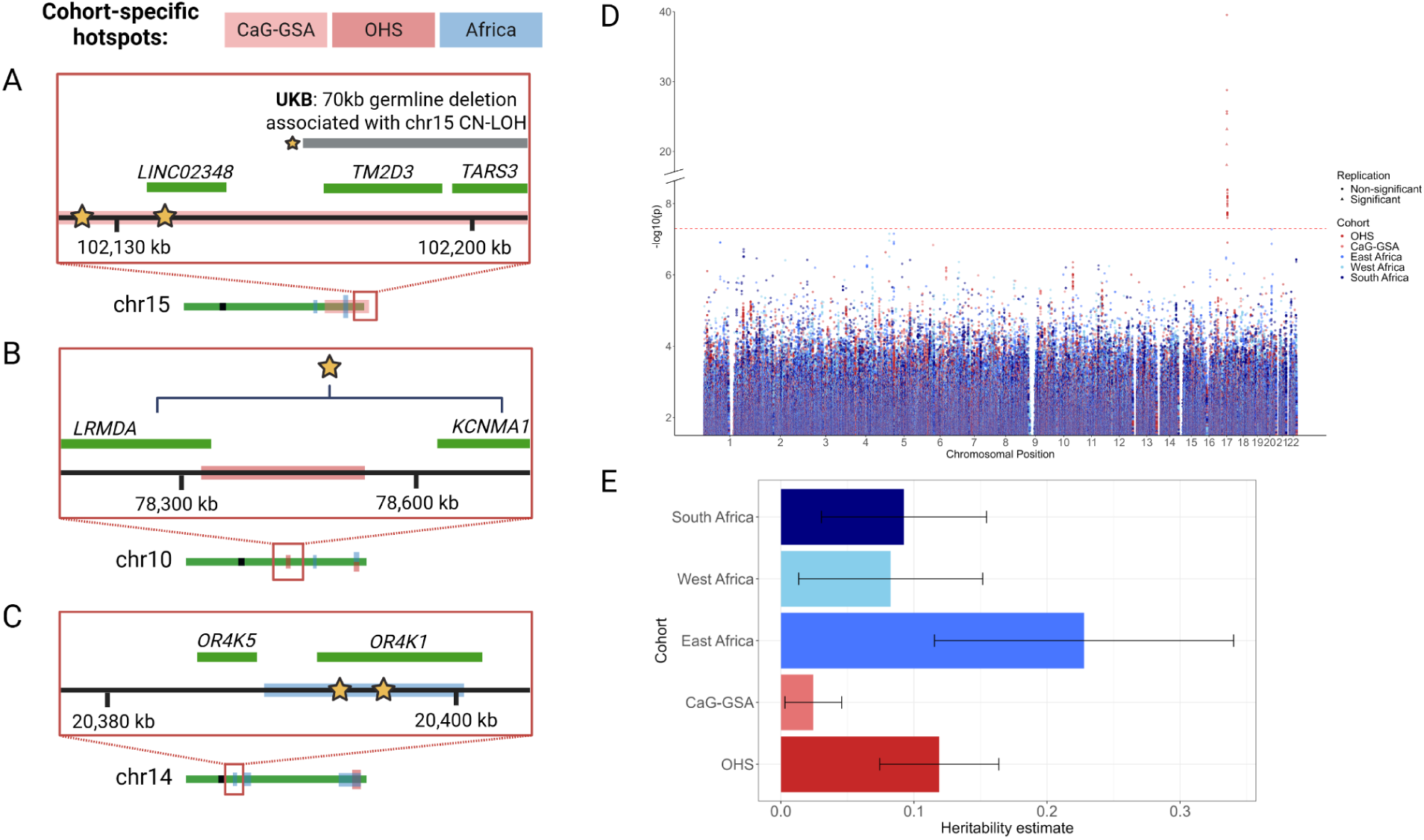
Germline genetic factors and heritability estimates associated with mCA prevalence. Graphical representation of germline variants associated with (A) CN-LOH mCAs in CaG-GSA, (B) CN-LOH and type-agnostic mCAs in OHS, and (C) type-agnostic mCAs in West Africa. The star symbols indicate approximate positions of hits, and brackets indicate the range containing multiple hits in linkage disequilibrium. The germline deletion associated with chr15 CN-LOH events in *cis* identified in the UK Biobank^11,12^ is highlighted with a star and in grey in (A). (D) Manhattan plot for autosome-wide associations with type-agnostic mCAs, in each cohort. The data points are colored by cohort. A genome-wide significance threshold of 5e-8 is indicated by the red dashed line. P-values in the discovery stage are plotted, with hits that were significant in the replication stage following linkage disequilibrium pruning indicated by triangular data points. (E) Heritability estimates for ‘type-agnostic’ autosomal mCA occurrence in each cohort.

**Table 1.**
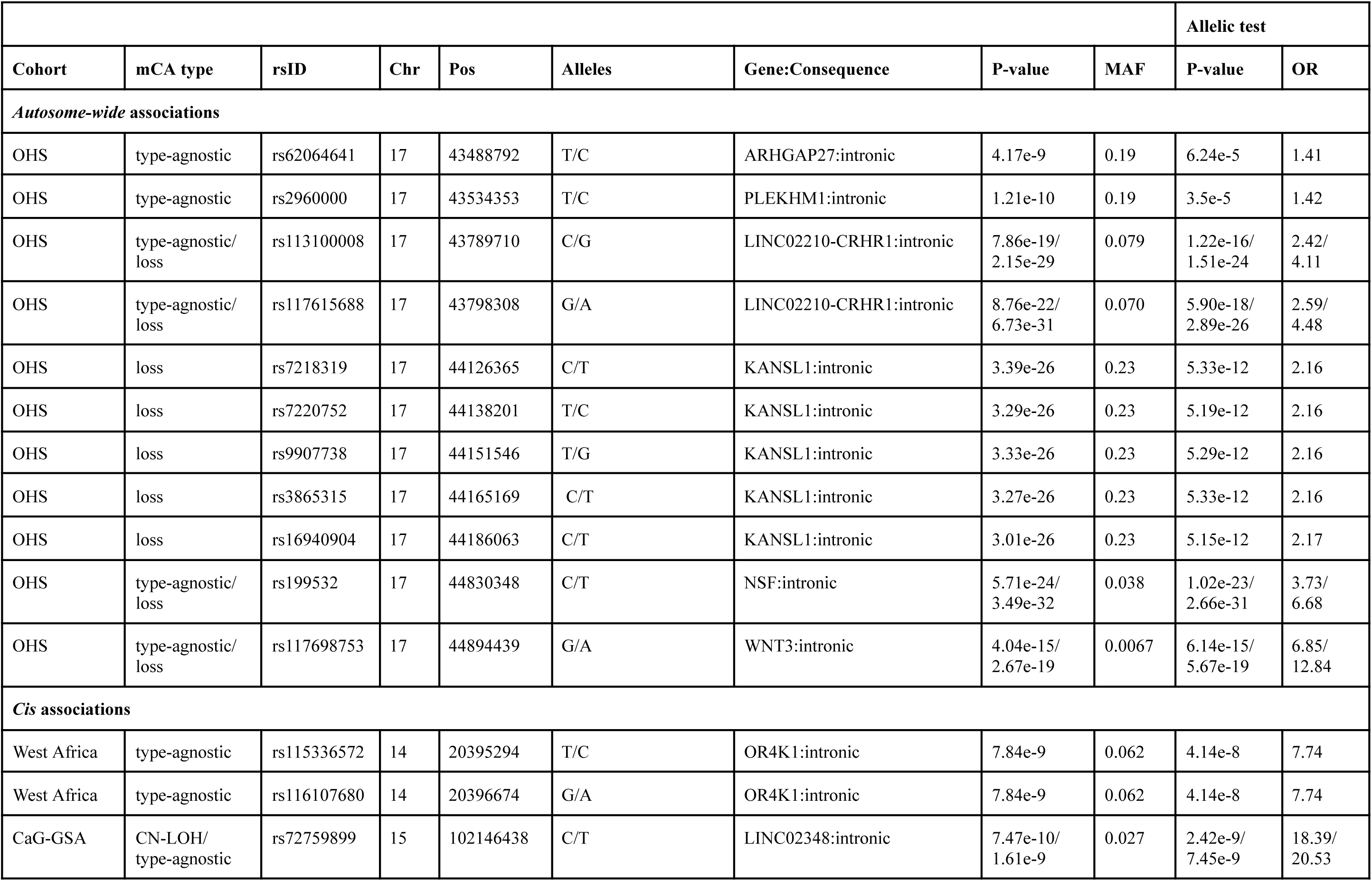
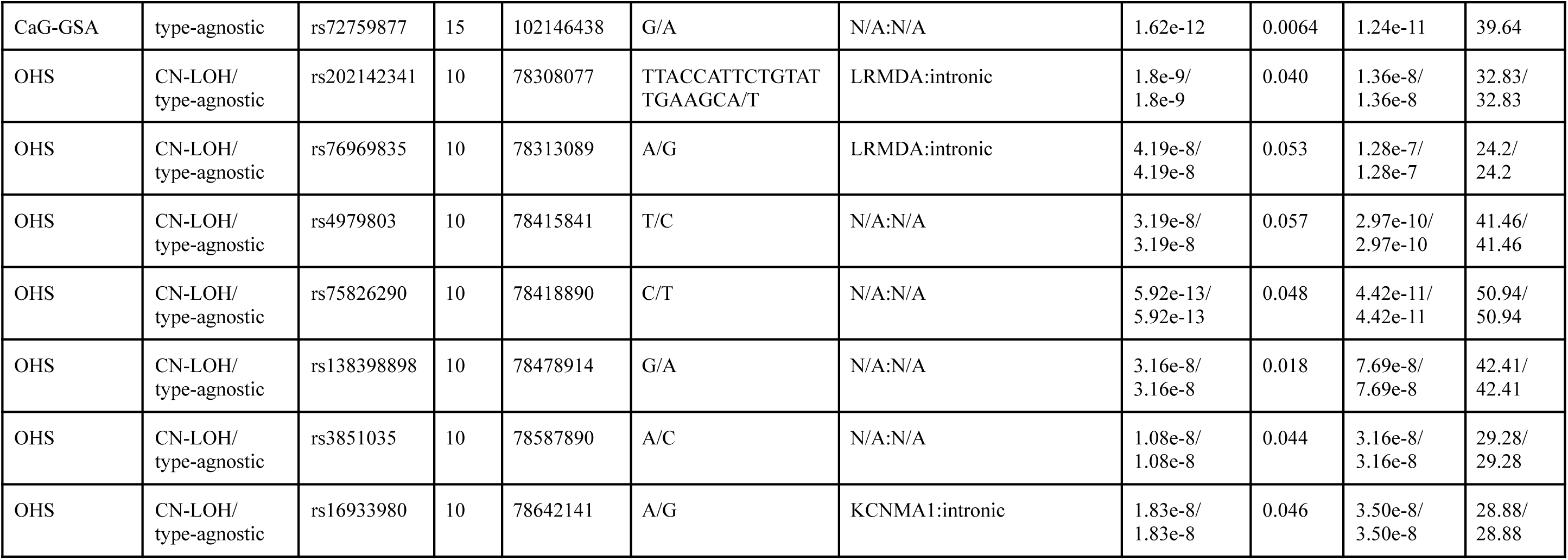
Germline variants in association with mCAs in *cis* or autosome-wide, in each cohort. Variants are specified by their association with a specific type of alteration (type-agnostic, gain, loss, CN-LOH). Base pair coordinates are mapped to hg19. “N/A” values in the Gene:Consequence field indicate that the variant did not map to a coding region. Minor allele frequencies (MAF) are reported for the variant within the cohort of discovery. When a variant is associated with more than one type of alteration, p-values and odds ratio (OR) are reported for each type of alteration, separated by a slash (“/”), in the order that the types are reported.

We next performed an association test to identify variants influencing autosome-wide mCA occurrence within every cohort, with the Bonferroni-corrected genome-wide significance threshold of 5.0e-8. Novel significant autosomal intronic variants within 17q21.31 were associated with autosomal type-agnostic (Figure 4D) and loss events within the Ontario cohort only (Table 1). Variants at 17q21.31 were located among the *LINC02210-CRHR1, NSF* and *WNT3* loci. Other variants at 17q21.31 associated uniquely with type-agnostic events were located in the *ARHGAP27* and *PLEKHM1* loci, while variants associated uniquely with loss events were located in the *KANSL1* locus.

Studies in various cohorts have reported multiple loci associated with mCA occurrence, such as, but not limited to: *ATM*^11,13,20^, *MPL*^11,13,20^, *FRA10B*^11^, *NBN*^11–13^, and *TCL1A*^13^ associated with mCAs in *cis*, and *MAD1L1*^13^, *TERT*^13,20^, and *TET2*^20^ associated with mCAs autosome-wide. We verified that we were able to observe many of these same variants as segregating in our North American cohorts yet despite their presence, associations were not significant, or did not reach the genome-wide significance threshold, perhaps due to smaller sample sizes in our cohorts. However, many of these previous mapped loci include variants that, in many cases, were not segregating in the African cohorts.

Given that individual germline segregating variants are mapping significantly to either mCA hotspots or autosome-wide mCA events, we next asked whether we could quantify the overall heritability of mCA accumulation with age. The population heritability (*h*^2^) estimate is defined as the proportion of variation in mCA occurrence that is attributable to genetic variants^54^. For continental African cohorts we observed similar autosomal heritability estimates and standard errors (*h*^2^ ∼ 8-22%), whereas in Quebec and Ontario we observed *h*^2^ estimates of 2.4% and 11.8%, respectively (Figure 4E). Estimates of *h*^2^ in East Africa were greater than other African cohorts, potentially suggesting that variants with larger effect sizes associated with autosome-wide mCA occurrence are present in the cohort, but not statistically significant in our analysis due to smaller sample sizes for the East African cohort. Previous heritability estimates for female loss of chromosome X were estimated to be 10.6% (s.e. 3.6%)^11^.

### mCAs impact the bulk blood transcriptome in a copy change-specific manner and confound estimates of allele-specific expression (ASE)

To determine the impact of mCAs on blood-based molecular phenotypes, we investigated the relationship between mCAs and the transcriptome available in the French-Canadian founding population (CaG-OMNI). While the interaction of somatic point mutations on tissue-specific transcriptomes has been studied among tumours^55^, the impact of mCAs on the healthy blood transcriptome has not been well-characterized and would significantly advance our understanding of why genomic structural perturbations are functionally tolerated at relatively high frequencies in blood.

To evaluate the global impact of mCAs on gene expression, we performed a burden-type analysis^56^ to examine the distribution of mCA types across the gene expression continuum. We limited our analysis to all transcripts which overlap with a detected autosomal mCA. All read counts were normalized to library size, scaled across each transcript, and adjusted by the proportion of lymphocytes in each sample (see Methods). Briefly, normalized expression values for each transcript were ranked across individuals and binned into expression percentiles. The number of mCAs overlapping a transcript were summed within each percentile bin iteratively across all transcripts. The cumulative burden of each type of mCA was then regressed across gene expression percentiles using separate linear and quadratic models and a permutation analysis was performed (n = 10,000) to determine if the observed burden of mCAs on gene expression was extreme (Figure 5A). Under neutrality, we would expect gene expression to be linearly correlated with copy number. We observed a significant excess of CN-LOHs (R^2^ = 0.68, p = 0.039, quadratic model) and a marginal excess of loss (R^2^ = 0.58, p = 0.055, quadratic model) events at extreme levels of gene expression. In contrast, we observed an excess of gain events at intermediate levels of gene expression across individuals (R^2^ = 0.67, p = 0.034, quadratic model). A linear model captured a significant association between gene expression and the burden of loss events (R^2^ = 0.19, p = 3.7e-3, linear model) but accounted for less variation in the association between mCAs and gene expression variability across participants compared to the quadratic model. Together, our findings suggest that gene expression stabilizes following a copy number gain but cannot recover from a copy number loss or loss of heterozygosity, likely due to the loss of the gene body or key regulatory regions.

**Fig. 5.**
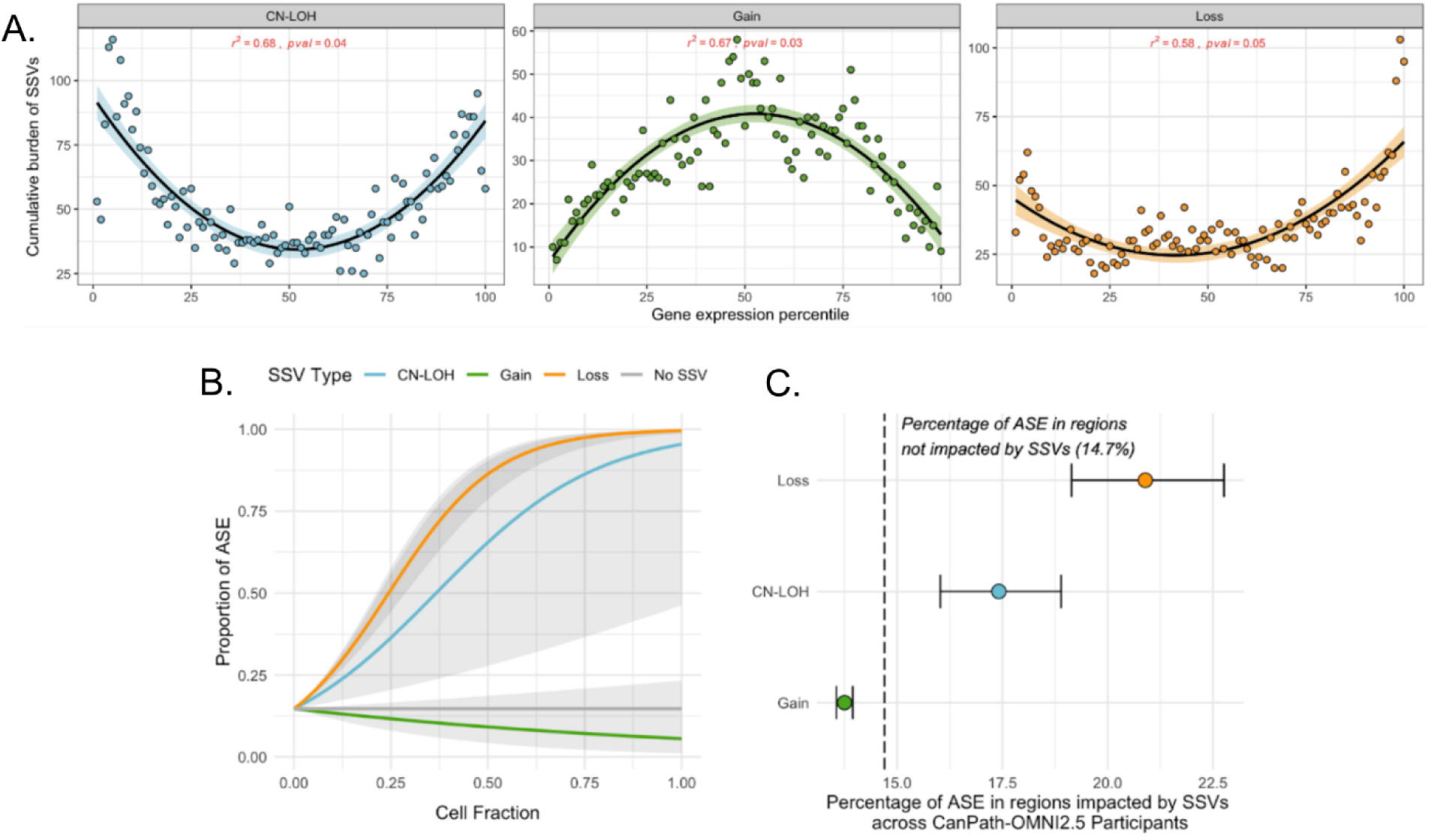
mCAs impact bulk blood transcriptomes in a subset of the French-Canadian founding population cohort. (A) Each point represents the cumulative burden of mCAs within a gene expression percentile bin, using the CaG-OMNI cohort in Quebec. A quadratic model is used to capture the association between the cumulative burden of mCAs and gene expression percentiles. Gain=green, Loss=orange, CN-LOH=blue. (B) The mean proportion of ASE is indicated by the coloured line and the shaded bars represent standard error of the mean. (C) The mean proportion of ASE is shown by each point and the error bars represent standard error of the mean. The dashed line indicates the proportion of ASE that is expected in those regions in the absence of mCAs (14.7%).

Finally, we investigated the impact of mCAs on the transcriptome by measuring allele-specific expression (ASE). ASE is a documented example of how selection can act at the transcriptional level to drive transcriptional variability at heterozygous regions, with genomic imprinting being the most extreme case^57^. ASE refers to the preferential expression of one of two parental alleles at a particular locus due to variations in regulatory sequences. We may expect mCAs to contribute towards estimates of ASE prevalence, due to the removal or duplication of alleles at loci.

To determine the impact of mCAs on estimates of ASE, we performed mCA-agnostic ASE calls in the French-Canadian founding population, as previously described^58^. Using a beta (zero and one) inflated regression, we modelled the proportion of sites with significant ASE as a function of cell fraction for each mCA type, as well as within regions not impacted by an mCA. As the degree to which ASE is confounded by mCAs is directly related to the frequency of each mCA in an individual’s blood, we estimated the proportion of sites exhibiting ASE across the cell fraction continuum (ranging from zero to one) for all mCA types (Figure 5B). We found that 14.7% of assessed sites in regions outside of mCAs exhibited ASE, consistent with previous estimates^59,60^ (Figure 5C). In contrast, the proportion of sites exhibiting ASE was higher in regions impacted by loss and CN-LOH mCAs, and lower in regions impacted by gain mCAs (Figure 5C).

## Discussion

Many studies in recent years have focused on how somatic mutations, which accumulate in our blood as we age, increase risk of progressing to cancer and cardiovascular disease^16,61,62^. Most of these studies have focused on the contributions of somatic point mutations and small insertions and deletions. Here we describe the contribution of ancestry, geography and sex on mosaic chromosomal alterations, and their impact on molecular phenotypes. Through leveraging a variety of genotyping studies in North America and continental Africa, we capture a consistent signature of how somatic mutations accumulate over the course of our lifespan. While SNP density has important implications for the capture of mCAs, mCA frequencies and genomic distributions in blood are shaped in part due to heritable factors and have an impact on molecular phenotypes such as gene expression, which may better link the impact of DNA structural changes arising in blood cells to disease risk.

Our approach to identify hotspots of mCAs across diverse ancestries offers a chromosome-specific view on where mCAs are more likely to be tolerated, and what temporal and population structures may underlie these patterns of accrual. The limitations of our approach lie in the assumptions made by the statistical model selected; namely, we assume a null mCA distribution as uniform across each chromosome and group mCAs of all types together. However, these assumptions may not align with the mechanisms of mCA formation and selection, as the probability of double-stranded breaks and DNA repair are impacted by sequence-level characteristics, and different mCA types are not formed at equal rates. Nonetheless, we adopted a pragmatic null model assuming uniform distribution across each chromosome which may point to these potential sources of population-specific variation. Our observations of continental or founding population specific differences may implicate genomic features that may segregate within populations, such as inversion events, that contribute to our observed variation. In addition, we present a large-scale harmonization effort to integrate array-based mCA calls from multiple cohorts in an unbiased manner, providing a methodological framework for future work in mCA characterization across genotyping platforms.

We report novel germline variants associated in *cis* and autosome-wide with mCAs in North American (Ontario, Quebec) and continental African (West Africa) cohorts, with some potentially having clinical relevance. Significant variants associated with autosome-wide events were only identified in the OHS cohort, in the 17q21.31 locus. Of the identified loci in this region, hits were found in the *WNT3* locus, a member of the Wnt family that is involved in the Wnt/ꞵ-catenin signaling pathway. The deregulation of this signaling pathway is known to be implicated in the progression of certain cancers^63,64^, including leukemias^65^, and chromosomal instability^66^, and therefore poses the possibility that the identified variant, or any SNPs within the locus that this variant may be tagging, may influence Wnt signaling regulation, providing a proliferative advantage to the clone. For *cis* associations, in Ontario, we identify significant variants within the 3’ UTR region of the *KCNMA1* locus, which is a tumor suppressor in gastric cancer^52^. While the *LINC02348* locus identified in the Quebec cohort has not been previously reported as being associated with mosaic events, it is located approximately 20 Kb upstream of the *TM2D3* gene, a locus which is overlapped by a rare germline deletion spanning to *TARS3* (70Kb deletion, tagged by rs182643535) associated with chr 15q CN-LOH and loss events in the UKB^11,12^. Therefore, it is possible that our variant identified in Quebec may be mapping this deletion. While the discovery and replication of *cis* associations in Ontario, Quebec, and West Africa was guided by cohort-specific hotspots identified from mCAs called with all assayed SNPs (Table S2), attempting to replicate associations across ancestries autosome-wide did not yield significant associations, suggesting that ancestry-specific germline variation shapes population-specific patterns of mCA acquisition. Alternatively, selection effects may be different across populations living in diverse environments. Additionally, none of the *cis* associations in the continental African cohorts, nor any variants approaching significance, were shared across at least two geographic regions (Table 1), and while hotspots are not unique to a geographic region within continental Africa, we show that germline variants associated with mCA occurrence at hotspots demonstrate region specificity within the continent.

The relative effects of ancestry, germline alleles, and environmental factors on the variance in mCAs across populations remain unclear. Our heritability analyses suggest that while some variation can be explained through genetic associations, much of the variation observed is either stochastic or explained by other factors such as age-associated environmental exposures. However, we were able to compare individuals of African ancestry in the North American cohorts (who were previously removed from analysis) with individuals of African ancestry in the continental African cohorts with respect to mCA accrual, to a limited extent due to sample sizes. Ten autosomal mCAs were found in Quebec and 4 in Ontario, for a total of 14 mCAs from individuals of African ancestry. Only 2/14 mCAs overlapped African cohort hotspots, and this discrepancy between continental and North American African hotspots was significant (p=0.043, one-tailed FET), suggesting that either geography or environment may impact the genomic location of mCA accrual. The limited sample size of individuals of African ancestry in the North American cohorts precludes any robust conclusions, but this observation could lay the groundwork for future studies to dissect the impact of genetic ancestry versus environment on mCA predisposition.

Previously, we characterized the impact of positive, negative and ‘interfering’ selection on the cellular distribution of point mutations^3^. We expect that mCAs are likely to be subject to a similar range of selection pressure due to the simultaneous disruption of multiple genes and regulatory elements along a chromosome. The observation that mCAs in hotspots have significantly reduced cell fractions relative to the rest of the genome implicates either negative selection, or potentially other undetected clones arising at lower frequencies due to driver mutations at these sites, and potentially interfering with each other’s clonal frequency, also known as Hill-Robertson interference^67^. A similar observation of reduced cell fraction with advanced age in many of our cohorts potentially corroborates that hypothesis. At the transcriptional level, we observe signatures of directional selection in the presence of loss and CN-LOH variants but find that gene expression stabilizes to a population average following gains, suggesting that there may be compensatory mechanisms acting at the transcriptomic level in the Quebec cohort to compensate for DNA level changes. Many studies have shown that gene expression levels are subject to stabilizing selection in model organisms, but to date it has not previously been observed in humans^68–70^.

Our study illustrates somatic autosomal mCA accrual similarities and distinctions across ancestries, geographies, and environments, and what germline factors and sex biases may underlie these differences. Future work using higher coverage or long-read sequencing will improve our understanding of the finer scale resolution of somatic structural variation in the hematopoietic pool that arise over our lifespan and their implications for disease development.

## Resource Availability

### Lead contact

Philip Awadalla;

### Materials availability

This study did not generate new unique reagents.

### Data and code availability

- Our data are accessible through the Ontario Health Study, CARTaGENE, and H3Africa upon request. To apply for access, contact (submit an application https://h3abionet.github.io/catalogue/3_requests.html). We obtained a publicly available, de-identified list of mCAs called in ∼480,000 UK Biobank participants^12^.
- All original code has been deposited and is publicly available here https://github.com/JasmineRWKang/mCAs-diverse-ancestries/tree/main
- Any additional information required to reanalyze the data reported in this paper is available from the lead contact upon request.

## Data Availability

Our data are accessible through the Ontario Health Study, CARTaGENE, and H3Africa upon request. To apply for access, contact (submit an application https://h3abionet.github.io/catalogue/3_requests.html). We obtained a publicly available, de-identified list of mCAs called in ∼480,000 UK Biobank participants.
All original code has been deposited and is publicly available here https://github.com/JasmineRWKang/mCAs-diverse-ancestries/tree/main
Any additional information required to reanalyze the data reported in this paper is available from the lead contact upon request.

https://h3abionet.github.io/catalogue/3_requests.html

https://www.ontariohealthstudy.ca/for-researchers/data-access-process/

https://cartagene.qc.ca/en/researchers/access-request.html

## Acknowledgments

We thank the participants of the CanPath and the H3Africa Consortium studies. The AWI-Gen Collaborative Centre was funded by the National Human Genome Research Institute (NHGRI), the National Institute of Environmental Health Sciences (NIEHS), of the National Institutes of Health (NIH) under award number U54HG006938. Q.M. was partially supported by a NIH/NCI Cancer Center Support Grant (P30 CA008748) and a Canada Artificial Intelligence chair from the Canadian Institute for Advanced Research. K.S. was partially supported by a Vector Institute Research Grant. The data and biosamples used for this research were made available by the OHS and CARTaGENE, with the financial support from the Canadian Partnership Against Cancer, Health Canada, the Ontario Institute for Cancer Research and the Government of OntarioThe views expressed herein represent the views of the Authors and do not necessarily represent the views of Canadian Partnership Against Cancer, Health Canada, the Ontario Institute for Cancer Research or the Government of Ontario.

## Author contributions

Conceptualization, P.A., M.J.F. and K.S.; Methodology, J.R.W.K., M.J.F., Y.J.K., and K.S.; Formal Analysis, J.R.W.K., Y.J.K., K.S., D.S., V.B., M.J.F., Q.M., and P.A.; Investigation, J.R.W.K., Y.J.K., K.S., V.B.; Resources and Data Curation, E.M., J.M., H.N., A.M., Z.L., M.R.; Writing - Original Draft, J.R.W.K., Y.J.K., K.S. and P.A; Writing - Review & Editing, J.R.W.K., Y.J.K., V.B., M.J.F., J.E., H.N., S.H., Z.L., M.R., Q.M., P.A.; Visualization, J.R.W.K., Y.J.K., K.S., V.B.; Supervision, Project Administration, and Funding Acquisition, P.A.

## Supplemental information

Document S1. Figures S1-S5 and Tables S4-S5.

Table S1. Excel file containing additional data too large to fit in a PDF, related to Figure 1.

Table S2. Excel file containing additional data too large to fit in a PDF, related to Figure 3.

Table S3. Excel file containing additional data too large to fit in a PDF, related to Figure 3.

## Methods

### Study population

Participants were recruited from the Ontario Health Study^30^ (OHS) and CARTaGENE^31^ (CaG), constituent population cohorts within the Canadian Partnership for Tomorrow’s Health^24^ (CanPath). A total of 3805 samples of non-European ancestry or admixed ancestry (of both European and non-European ancestry) were removed from OHS and CaG. Participants of African ancestry were recruited from the Africa Wits-INDEPTH Partnership for Genomic Research^32^ (AWI-Gen) and TrypanoGEN+^34^ cohorts within the H3Africa consortium^21^. TrypanoGEN+ is composed of two sub-cohorts encompassing individuals with neglected tropical diseases, Trypanosomiasis and Schistosomiasis. AWI-Gen was established to examine genomic, environmental and behavioral factors influencing body composition and cardiometabolic diseases and traits in continental African populations, and include participants from Burkina Faso, Ghana, Kenya and South Africa. All participants provided broad consent for their data and biologics to be made available to the research community at the time of recruitment. No statistical methods were used to pre-determine sample size. The study protocol was approved by the Health Sciences Ethics Review Board Committee of the University of Toronto. Analyses involving mCA calls from ∼500,000 UKB participants were made using a publicly available dataset published by Loh et al^12^. 94.6% of the participants in the UKB are of white ethnicity, reflecting the national population at the time of recruitment^35^.

### Genotyping

One set of the genotyping data from CARTaGENE used in this study has been previously published by Favé et al.^71^, Hussin et al.^27^, Hodgkinson et al.^72^, and Harwood et al.^58^. Briefly, 937 samples with RNA-Seq profiles that passed quality control (QC) thresholds were genotyped on the Illumina OMNI2.5 array to obtain high-density SNP genotyping data. A second dataset of 30,269 samples from CARTaGENE (including 925 samples previously genotyped with OMNI2.5) were genotyped with the Infinium Global Screening Array (GSA) from Illumina. A total of 21,076 samples from the Ontario Health Study (OHS) were genotyped with the UKBiobank Axiom array. Finally, 13,562 samples of African ancestry from AWI-Gen and TrypanoGEN+, genotyped on the H3Africa array, have been used in this study. For all arrays, only genotypes with high quality were retained for further analyses (SNPs with call rate >= 97% and Hardy-Weinberg equilibrium (HWE) p-value > 1e-3).

### Population structure and ancestry inference

We used the Kinship-based INference for GWAS (KING 2.3.1) software^73^ to assess relatedness between all pairs of samples within each cohort. Pairs identified as second-degree relatives or closer were flagged. Population structure was computed by principal component analysis (PCA) using KING, applied to unrelated samples and LD-pruned SNPs. PCA was performed jointly for OHS, CaG, AWI-Gen and TrypanoGEN+. In addition to the principal components, KING can also infer the most likely ancestral group(s) for each sample, by leveraging known ancestry in a reference dataset, such as the 1000 Genomes Project data. It classifies samples into one or more of the following superpopulation groups: AFR (African), AMR (American), EAS (East Asian), EUR (European), and SAS (South Asian). This analysis showed that over 90% of samples from OHS and CARTaGENE cohorts have European ancestry. The remaining samples were roughly evenly split among African, American and Asian ancestry (Figure S1A). For the AWI-Gen and TrypanoGEN+ cohorts, ancestry inference confirmed that all individuals have African ancestry. The PCA plot focusing on these samples revealed distinct clustering by country, highlighting the continental African population structure (Figure S1B,C), with a high degree of heterogeneity seen in East Africa.

### Mosaic chromosomal alteration (mCA) calling

All mosaic chromosomal alteration calling was performed using the publicly available MoChA v1.11 software^11^. This approach utilizes phased genotypes, log R ratio (LRR) and B allele frequency (BAF) (which measure total and relative allelic intensities, respectively) at heterozygous sites for detection of mCAs. For each array separately, SNPs within 2 s.d. of the mean LRR and BAF distribution were selected. As such, SNPs with poor genotype quality or increased intensity noise were excluded. Genotypes were phased using EAGLE2^74^ with the Haplotype reference Consortium panel for CARTaGENE and OHS, and the African Genome Resources panel for AWI-Gen and TrypanoGEN+ cohorts. As recommended by the author of MoChA, variants falling within segmental duplications from the UCSC genome annotation database with low divergence (Jukes–Cantor distance < 2%), high levels of missingness (>3%) and excess heterozygosity (p<1e-6) were excluded. For all arrays, the default parameters for the HMM model were applied, the common germline duplications and deletions from 1000 Genomes phase3, the MHC and KIR regions were excluded from the analysis as suggested by the developer. Calls were filtered according to the default MoChA suggestion. Briefly, we removed samples with call-rates lower than 0.97 and a BAF-auto greater than 0.03. Additionally, we removed calls made by the LRR and BAF model if they had a lod_baf_phase score less than 10 for the model based on BAF and genotype phase. Individuals genotyped using the OMNI2.5 and GSA array were mapped to hg19. Marker coordinates for individuals genotyped using the UKBiobank Axiom Array (hg38) were converted to hg19 using *liftover*^75^. mCAs were either called using all retained SNPs on a given array or SNPs downsampled to the least dense array (GSA and Axiom, ∼600,000 SNPs). To downsample SNPs in H3Africa, sites were selected such that the pairwise distance between any two sites was greater than 2kb, reflecting the density of GSA and Axiom arrays. We removed mCA calls from the region from 43-45Mb on chr 17 in OHS from our analyses, due to the known structural complexity of the 17q21.31 locus and the existence of two copy number polymorphic segmental duplications, which are flagged as inherited in multiple genomic databases^76^.

### Identification of mCA hotspots

mCA hotspots were identified in each cohort using a right-tailed binomial test approach, querying start and stop sites of mCAs on each autosome. The total length queried on each chromosome (distance from the first to last SNP on the array) and a list of mCA start/stop sites occurring on the chromosome was constructed for each cohort. The number of overlapping mCAs at each site, the total number of mCAs on the chromosome, and the chromosome-specific baseline levels of mCA burden were used to conduct a right-tailed binomial test at each site. Chromosome-specific baseline levels of mCA burden were calculated as the average mCA length on the chromosome divided by the total chromosomal length queried. The mean mCA length differed considerably across chromosomes and across cohort-arrays, which was reflected in the variance in chromosome-specific baseline levels of mCA burden. A Benjamini-Hochberg FDR-corrected threshold was calculated using the total number of binomial tests conducted. mCAs of undetermined copy change type and those occurring on sex chromosomes were discarded.

### mCA hotspot region overlaps

mCA hotspot regions were defined as continuous genomic stretches in which every queried site within the region had a binomial test surpassing the BH-corrected significance threshold. Two regions were defined as separate if the SNPs at adjacent ends of the regions were at least 15 Mb apart. For cross-cohort comparisons, hotspot regions were identified from mCAs called with SNPs downsampled to the least-dense arrays. The degree of overlap between hotspot regions was calculated for every possible cohort pair using the GenomicRanges R package^77^ (v1.58.0). To compare hotspot regions between sexes of the same cohort, hotspot regions were identified from mCAs called with all SNPs in the given array and GenomicRanges was similarly used to quantify percent overlap, ie. the length of the overlap divided by the total length of the hotspot region.

### Association of germline variants with cis and autosome-wide mCA events

Several related pairs from the first-to fourth-degree were identified within each of our ancestry-specific cohorts using KING^73^, as previously stated. Familial relationships were inferred from pairs related up to the second degree, and each familial unit was pruned to keep one sample, with a preference to keep a sample if it had a mosaic chromosomal event. The association analyses were thus restricted to 16,093 OHS, 25,750 CaG-GSA, and 11,834 African (pooled AWI-Gen and TrypanoGEN+) unrelated samples.

Continental African populations have a high degree of genetic diversity, which can confound genetic association studies^22,23,39,78^. To account for the population substructure within the continental African population cohorts, we stratified samples from AWI-Gen and TrypanoGEN+ into geographical regions of the continent (East, South, West, and Central) based on the sample’s country of origin^23^ and conducted genetic association tests on each geographical cohort separately (Table S5). Samples that did not have any available data on the country of origin were excluded from association analyses, resulting in a final continental African cohort sample size of 11,372 across all geographical regions.

Imputation was performed on the TOPMED Imputation Server^79^ (Minimac3), using the TOPMED-r3 reference panel for the European ancestry cohorts and the African Genome Resources panel for the African-ancestry cohorts. All imputed variants were filtered to R^2^>=0.4, a minimum minor allele frequency of 0.5% and variant missing call rate within the cohort below 2%. Approximately 8.5 million and 7.8 million imputed variants passed these quality control filters in the European ancestry cohorts CaG-GSA and OHS, respectively. In the continental African cohorts, around 13 to 15 million imputed variants passed these quality control filters for each geographical cohort. The Bonferroni correction procedure was performed for each association analysis.

We performed a chromosome-wide association analysis to identify inherited autosomal variants influencing somatic mosaic chromosomal events overlapping hotspots in *cis*, following the methodology described by Loh et al^11^. Cases are characterized on a per-variant basis, defined as a sample with a hotspot-overlapping mosaic event present nearby (within a 4Mbp range) or overlapping the variant. To minimize the burden of multiple testing, variants were filtered based on the number of cases, with a minimum threshold of 5 and 3 set for the CaG-GSA and OHS cohorts respectively, and a threshold of 2 for CaG-OMNI and each continental African geographic cohort. These threshold values were extrapolated from the threshold of 25 used by Loh et al^11^ to be scaled to our cohort sizes. Fisher’s exact test was conducted to identify associations between variants and autosomal mosaic chromosomal events of any type, and events stratified by alteration type (gain, loss, CN-LOH, type-agnostic).

An autosome-wide association analysis was performed to identify autosomal germline variants influencing the occurrence of autosome-wide mosaic events. mCA phenotypes are characterized by the type of alteration (gain, loss, CN-LOH, type-agnostic) occurring on an autosome. The autosome-wide association testing was performed using PLINK 1.9^80^, with a genome-wide significance threshold of 5.0e-8.

### Heritability analysis of autosomal mCAs

Quality control thresholds for imputation quality, minor allele frequency and genotype missingness were identical to those used in the association analyses. Following these filters, linkage disequilibrium pruning was performed on each cohort until approximately 500,000 SNPs remained. BOLT-REML^81^ was used to estimate heritability estimates and standard errors.

### RNA sequencing

RNA sequencing data from CaG-OMNI have been previously published by Favé et al.^71^, Hussin et al.^27^, Hodgkinson et al.^72^, and Harwood et al.^58^ Briefly, whole blood samples were collected from participants in 2010. Total RNA was isolated using the Tempus Spin RNA isolation kit (ThermoFisher Scientific) and a globin mRNA-depletion was performed using the GLOBINclear-Human kit (ThermoFisher Scientific). TruSeq RNA Sample Prep kit v2 (Illumina) was used to construct paired-end RNA-Seq libraries with 500 ng of globin-depleted total RNA. Recommended Illumina protocols were followed for quantification and quality control of RNASeq libraries prior to sequencing. Paired-end RNA sequencing was performed on a HiSeq 2000 platform at the Genome Quebec Innovation Center (Montreal, Canada). Sixty million reads per sample were captured. All RNA-seq experimental steps following blood draw were conducted in the same central laboratory, and samples were distributed randomly over sequencing lanes, thereby reducing the introduction of experimental bias at these steps. The RNA-Seq data was processed and prepared as described in Favé et al^71^, using counts from HTSeq and upon which we applied an inverse normal transformation.

### Normalization of CanPath Expression data

Raw expression counts were available for 17,224 transcripts across 997 individuals in the CaG-OMNI dataset. After filtering for transcripts which overlap with at least one mosaic chromosomal alteration, we retained 6,763 for downstream analyses. Expression counts were normalized by library size and proportion of lymphocytes in each sample. 151 individuals where lymphocyte proportions were missing were removed and we retained expression values for 846 individuals. Subsequently, the normalized counts were scaled across each transcript.

### Burden analysis

An approach described previously^56^ was modified to test if somatic mosaicism impacts gene expression. The range of gene expression was evaluated across 6,763 transcripts for 846 individuals with a total of 231 mosaic chromosomal alterations. Normalized expression values for each transcript were ranked across individuals and binned into 100 equal sized expression percentiles. The number of mCAs that overlap a transcript were summed within percentile bins across all transcripts. The cumulative sum of mosaic chromosomal alterations was regressed on expression percentiles using separate linear and quadratic models for each variant type: gain, loss and CN-LOH. The departure from the null expectation was assessed by evaluating the significance of the quadratic model relative to 10,000 permutations of the gene expression matrix.

### Modeling the impact of mCAs on estimates of allele specific expression

Allele specific expression was assessed as previously described^58^. Briefly, ASE estimates were calculated for germline heterozygous positions captured in the CaG-OMNI dataset. Significant ASE sites were determined using two-tailed binomial tests with Benjamini-Hochberg multiple testing correction (FDR < 0.05) to test the null hypothesis that *P* = 0.5, where *P* is the proportion of reads with the derived allele divided by the total read count for the site. For the 713,843 individual-SNP pairs, significant ASE was separated into overexpression and underexpression of the alternative allele by dividing the alternative allele count by the total read count. An alternative allele proportion of >0.5 demonstrates overexpression of the alternative allele and <0.5 demonstrates underexpression of the alternative allele. We used zero- and one-inflated beta regression to model the proportion of sites with significant ASE as a function of cell fraction for each mCA type (gains, losses and CN-LOH), as well as within regions not impacted by somatic mosaicism. Random effects were included to account for repeated observations within individuals. Beta regression modeling was performed using the gamlss R package (v5.3.4).

### Analytical environment and packages used

Analyses and figures were performed in the R statistical programming environment (v4.4.3). Libraries that are required include tidyr v1.3.1, purrr v1.0.4, plyr v1.8.6, dplyr v1.0.5, stringr v1.4.0, data.table v1.14.0, binom v1.1.1.1, betareg v3.2.2, marginaleffects v0.25.1, biomaRt v2.46.3, GenomicRanges v1.58.0, karyoploteR v1.32.0, survival v3.8.3, survminer v0.5.0, lubridate v1.9.4, gamlss v5.3-4, Hmisc v4.5-0, plotrix v3.8-1, ggeffects v1.1.2.1, igraph v2.1.4, foreach v1.5.2, and doParallel v1.0.17. Figures were generated using ggplot2 v3.3.3, ggsignif v0.6.4, ggpubr v040, ggplotify v0.0.7, and patchwork v1.1.1. All Wilcoxon rank-sum tests are two-sided unless otherwise specified.

