## Supplementary Figures and Tables S4, S5 for "Geography, Ancestry, Age and Sex Shape Somatic Autosomal Mosaic Chromosomal Alterations in Blood"

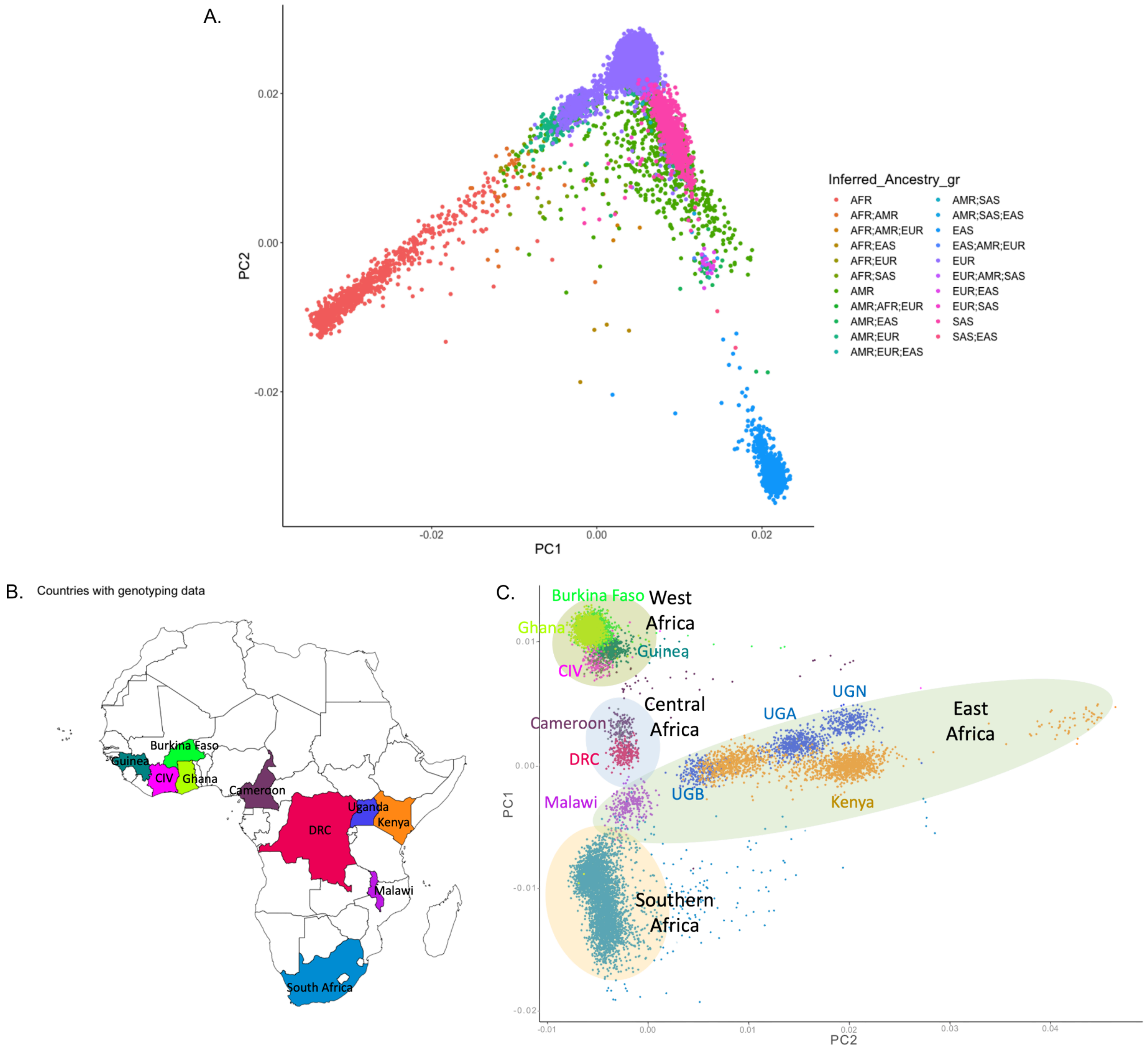


**Fig. S1. Population structure and inferred ancestry in North American and continental African cohorts.** (A) Principal component analysis of samples in North American cohorts (OHS, CaG-Omni, CaG-GSA), with inferred ancestry labels from 1000 Genomes Project. (B) Map of African countries sampled by AWI-Gen and TrypanoGEN+. (C) Principal component analysis of all samples in the continental African cohorts (AWI-Gen, TrypanoGEN+), which are of African ancestry. Black text indicates labels of geographic regions (West, East, Central, and Southern Africa). OHS=Ontario Health Study, CaG=CARTaGENE, GSA=Global Screening Array, AWI-Gen=Africa Wits-INDEPTH Partnership for Genomic Research.

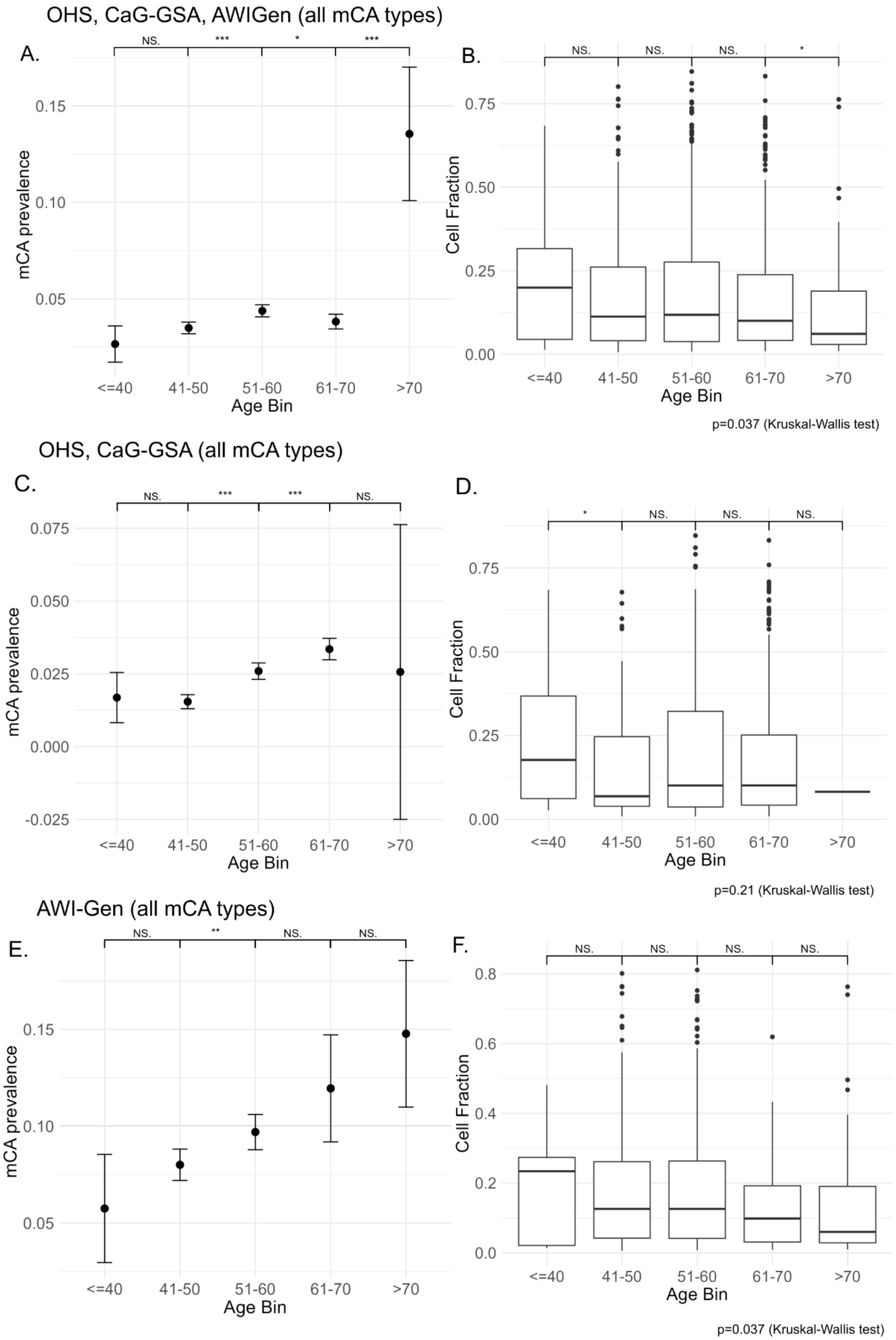


**Fig. S2. Characterization of mCA prevalence and cell fraction by age.** (A, B) Proportion of samples harboring at least one mCA and cell fractions of mCAs across age in OHS, CaG-GSA, and AWI-Gen. (C, D) Proportion of samples harboring at least one mCA and cell fractions of mCAs across age in North American cohorts. (E, F) Proportion of samples harboring at least one mCA and cell fractions of mCAs across age in AWI-Gen. mCAs were called using all assayed SNPs and included all types (including undetermined copy number change). Samples of non-European ancestry in OHS and CaG were removed. Two-sample proportion tests were conducted in A, C, and E; dots represent proportions, bars represent 2 standard deviations. Wilcoxon rank-sum tests and Kruskal-Wallis tests were conducted in B, D, and F; lines represent medians, box heights represent interquartile ranges. * $p<0.05$, ** $p<0.01$, *** $p<0.001$. OHS=Ontario Health Study, CaG=CARTaGENE, GSA=Global Screening Array.

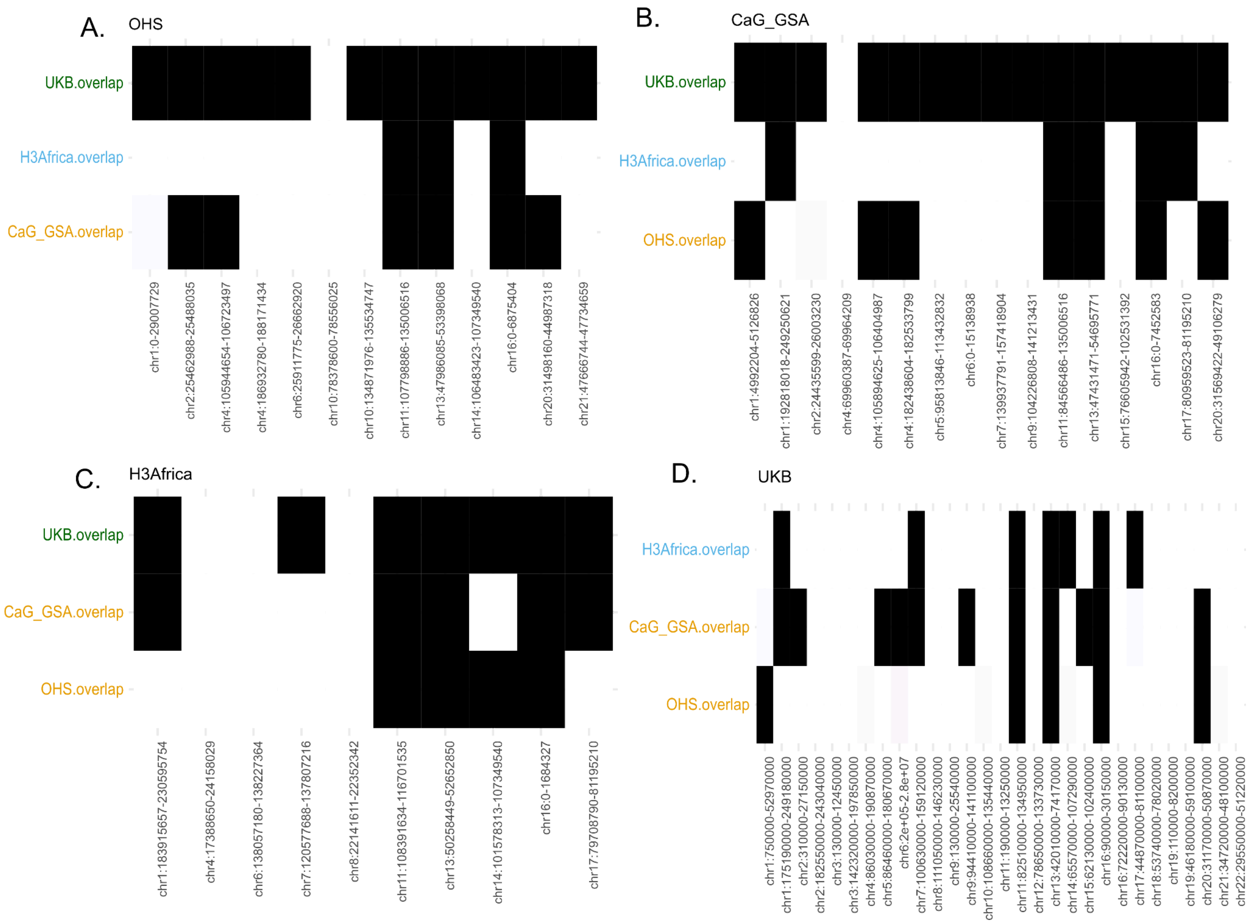


**Fig. S3. Autosomal mCA hotspots vary across populations and continents, related to Fig. 3.** mCAs were called from downsampled SNPs matched to the density of the least-dense arrays. Each SNP within a region surpasses the FDR-corrected threshold for significance in the binomial test. Each hotspot region's genomic coordinates in hg19 are listed on the x-axis for OHS-Axiom (A), CaG-GSA (B), H3Africa (C), and UKB-Axiom (D). Overlapping cohorts (y-axis) are coloured according to geographic region (orange=North America, blue=continental Africa, green=Europe). Filled black boxes indicate hotspot region overlaps, while unfilled boxes indicate hotspot regions unique to a given cohort (no overlap with any hotspot region in any other cohort). All samples of non-European ancestry were removed from OHS and CaG. Hotspots were called using BH multiple testing correction accounting for all binomial tests in OHS, CaG-GSA, H3Africa, and UKB (adjusted α = 0.023). OHS=Ontario Health Study, CaG=CARTaGENE, UKB=UK Biobank, GSA=Global Screening Array.

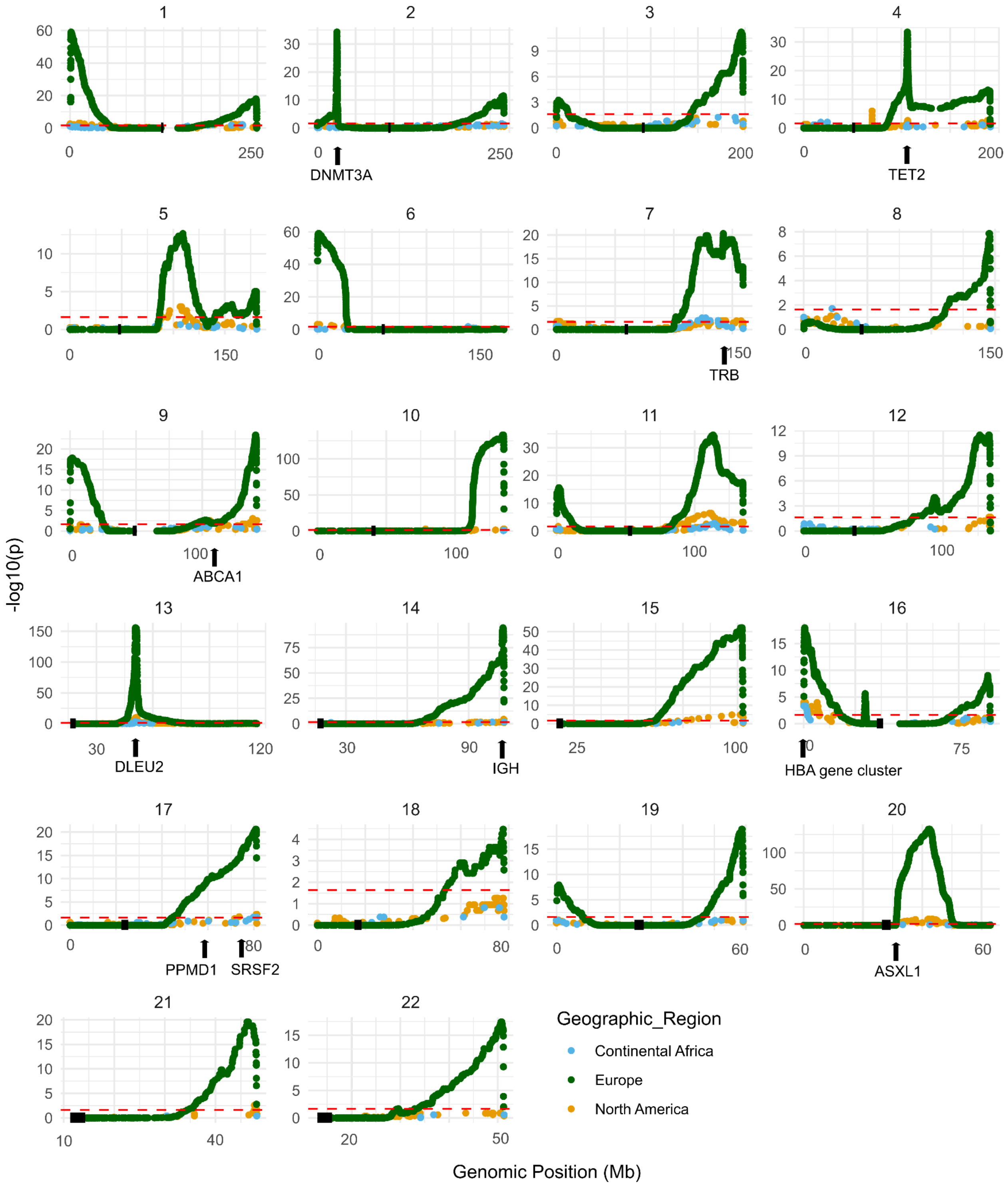


**Fig. S4. mCA hotspots identified in North American, continental African, and European cohorts, related to Fig. 3.** mCAs were called using downsampled SNPs to match the least dense arrays. Basepair coordinates on the x-axis are mapped to hg19. Centromeric regions in each chromosome are indicated in solid black. The significance threshold for hotspot identification after Benjamini-Hochberg multiple testing correction is shown as a red dotted line, (adjusted α = 0.023), adjusting for all binomial tests in North America, Europe, and continental Africa.

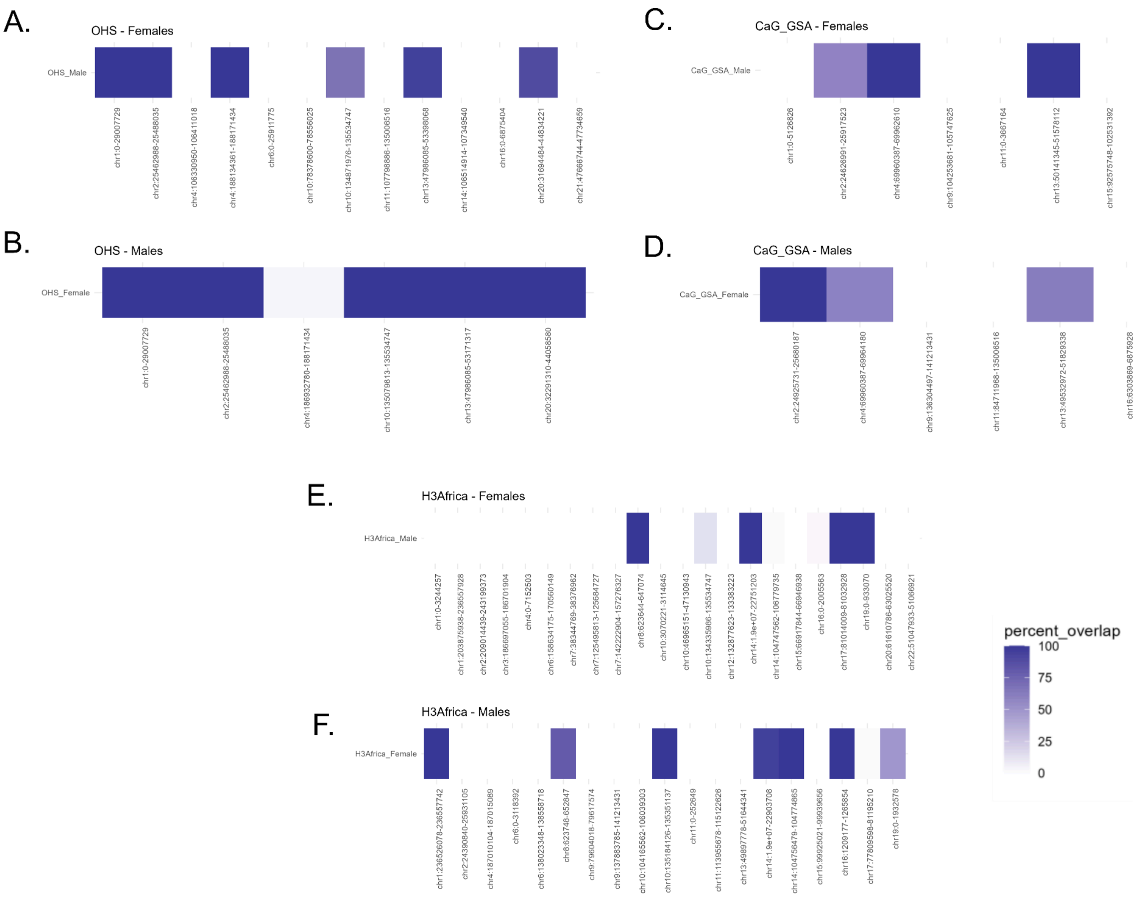


**Fig. S5. Autosomal mCA hotspots demonstrate sex specificity that is also geographically unique.** Female hotspot regions in (A) OHS, (C) CaG GSA, and (E) H3Africa; male hotspot regions in (B) OHS, (D) CaG GSA, and (F) H3Africa. Hotspot regions were identified separately in females and males for all cohorts, and annotated by degree of overlap with hotspot regions (h19 coordinates) found in the opposite sex. Each SNP within a hotspot region surpasses the FDR-corrected threshold for significance in the binomial test. Adjusted α = 9.4e-3 in females and 8.61e-3 in males, after Benjamini-Hochberg multiple testing correction accounting for all binomial tests in OHS, CaG GSA, and H3Africa in a given sex. Percent overlap of each hotspot region is quantified as the length of the overlapping region divided by the total length of the hotspot region. mCAs were called using all available SNPs (no downsampling). Each hotspot region's genomic coordinates are listed on the x-axis. All samples of non-European ancestry were removed from OHS and CaG. OHS=Ontario Health Study, CaG=CARTaGENE, GSA=Global Screening Array, UKB=UK Biobank.

**Table S1:** mCAs called from GSA and OMNI arrays (attached Excel file)

**Table S2:** Genomic positions of hotspot sites from mCAs called with all SNPs (attached Excel file)

**Table S3:** Genomic positions of hotspot sites from mCAs called with downsampled SNPs (attached Excel file), related to Fig. 3.

| Cohort-Array | Geographic Region | Chromosome | Start of hotspot region | End of hotspot region |
|---|---|---|---|---|
| OHS-Axiom | North America | 1 | 0 | 29007729 |
| OHS-Axiom | North America | 2 | 25462988 | 25488035 |
| OHS-Axiom | North America | 4 | 105944654 | 106723497 |
| OHS-Axiom | North America | 4 | 186932780 | 188171434 |
| OHS-Axiom | North America | 6 | 25911775 | 26662920 |
| OHS-Axiom | North America | 10 | 78378600 | 78556025 |
| OHS-Axiom | North America | 10 | 134871976 | 135534747 |
| OHS-Axiom | North America | 11 | 107798886 | 135006516 |
| OHS-Axiom | North America | 13 | 47986085 | 53398068 |
| OHS-Axiom | North America | 14 | 106483423 | 107349540 |
| OHS-Axiom | North America | 16 | 0 | 6875404 |
| OHS-Axiom | North America | 20 | 31498160 | 44987318 |
| OHS-Axiom | North America | 21 | 47666744 | 47734659 |
| CaG-GSA | North America | 1 | 4992204 | 5126826 |
| CaG-GSA | North America | 1 | 192818018 | 249250621 |
| CaG-GSA | North America | 2 | 24435599 | 26003230 |
| CaG-GSA | North America | 4 | 69960387 | 69964209 |
| CaG-GSA | North America | 4 | 105894625 | 106404987 |

| | | | | |
|---|---|---|---|---|
| CaG-GSA | North America | 4 | 182438604 | 182533799 |
| CaG-GSA | North America | 5 | 95813846 | 113432832 |
| CaG-GSA | North America | 6 | 0 | 15138938 |
| CaG-GSA | North America | 7 | 139937791 | 157418904 |
| CaG-GSA | North America | 9 | 104226808 | 141213431 |
| CaG-GSA | North America | 11 | 84566486 | 135006516 |
| CaG-GSA | North America | 13 | 47431471 | 54695771 |
| CaG-GSA | North America | 15 | 76605942 | 102531392 |
| CaG-GSA | North America | 16 | 0 | 7452583 |
| CaG-GSA | North America | 17 | 80959523 | 81195210 |
| CaG-GSA | North America | 20 | 31569422 | 49106279 |
| AWI-Gen/TrypanoGEN-H3Africa | Africa | 1 | 183915657 | 230595754 |
| AWI-Gen/TrypanoGEN-H3Africa | Africa | 4 | 17388650 | 24158029 |
| AWI-Gen/TrypanoGEN-H3Africa | Africa | 6 | 138057180 | 138227364 |
| AWI-Gen/TrypanoGEN-H3Africa | Africa | 7 | 120577688 | 137807216 |
| AWI-Gen/TrypanoGEN-H3Africa | Africa | 8 | 22141611 | 22352342 |
| AWI-Gen/TrypanoGEN-H3Africa | Africa | 11 | 108391634 | 116701535 |
| AWI-Gen/TrypanoGEN-H3Africa | Africa | 13 | 50258449 | 52652850 |
| AWI-Gen/TrypanoGEN-H3Africa | Africa | 14 | 101578313 | 107349540 |
| AWI-Gen/TrypanoGEN-H3Africa | Africa | 16 | 0 | 1684327 |
| AWI-Gen/TrypanoGEN-H3Africa | Africa | 17 | 79708790 | 81195210 |

| | | | | |
|---|---|---|---|---|
| UKB-Axiom | Europe | 1 | 750000 | 52970000 |
| UKB-Axiom | Europe | 1 | 175190000 | 249180000 |
| UKB-Axiom | Europe | 2 | 310000 | 27150000 |
| UKB-Axiom | Europe | 2 | 182550000 | 243040000 |
| UKB-Axiom | Europe | 3 | 130000 | 12450000 |
| UKB-Axiom | Europe | 3 | 142320000 | 197850000 |
| UKB-Axiom | Europe | 4 | 86030000 | 190870000 |
| UKB-Axiom | Europe | 5 | 86460000 | 180670000 |
| UKB-Axiom | Europe | 6 | 200000 | 28000000 |
| UKB-Axiom | Europe | 7 | 100630000 | 159120000 |
| UKB-Axiom | Europe | 8 | 111050000 | 146230000 |
| UKB-Axiom | Europe | 9 | 130000 | 25540000 |
| UKB-Axiom | Europe | 9 | 94410000 | 141100000 |
| UKB-Axiom | Europe | 10 | 108660000 | 135440000 |
| UKB-Axiom | Europe | 11 | 190000 | 13250000 |
| UKB-Axiom | Europe | 11 | 82510000 | 134950000 |
| UKB-Axiom | Europe | 12 | 78650000 | 133730000 |
| UKB-Axiom | Europe | 13 | 42010000 | 74170000 |
| UKB-Axiom | Europe | 14 | 65570000 | 107290000 |
| UKB-Axiom | Europe | 15 | 62130000 | 102400000 |
| UKB-Axiom | Europe | 16 | 90000 | 30150000 |
| UKB-Axiom | Europe | 16 | 72220000 | 90130000 |
| UKB-Axiom | Europe | 17 | 44870000 | 81100000 |

| | | | | |
|---|---|---|---|---|
| UKB-Axiom | Europe | 18 | 53740000 | 78020000 |
| UKB-Axiom | Europe | 19 | 110000 | 8200000 |
| UKB-Axiom | Europe | 19 | 46180000 | 59100000 |
| UKB-Axiom | Europe | 20 | 31170000 | 50870000 |
| UKB-Axiom | Europe | 21 | 34720000 | 48100000 |
| UKB-Axiom | Europe | 22 | 29550000 | 51220000 |

**Table S4: Hotspot regions in North American, continental African, and European cohorts.** Hotspot regions were defined as regions in which a binomial test at each SNP within the region surpassed FDR-corrected significance. mCAs were called using SNPs downsampled to match the least-dense array. Calls of undetermined copy number change were removed. Samples of non-European ancestry were removed from North American cohorts (OHS and CaG). Basepair start and stop positions are mapped to hg19. BH multiple testing correction accounted for all binomial tests conducted in OHS, CaG-GSA, H3Africa, and UKB (adjusted $\alpha = 0.023$).

| Country | N | Samples with autosomal mCA |
|---|---|---|
| **West** | | |
| Burkina Faso | 1775 | 130 |
| Cote d'Ivoire | 199 | 9 |
| Ghana | 1713 | 162 |
| Guinea | 587 | 21 |
| *Total* | 4274 | 322 |
| **East** | | |
| Kenya | 1756 | 119 |
| Malawi | 318 | 30 |
| Uganda | 729 | 72 |
| *Total* | 2803 | 221 |
| **Central** | | |
| Cameroon | 410 | 15 |
| Democratic Republic of the Congo | 262 | 16 |
| *Total* | 672 | 31 |
| **South** | | |
| South Africa | 4681 | 316 |
| *Total* | 4681 | 316 |
| **N.A.** | | |
| *Total* | 581 | 30 |

**Table S5: Stratification of samples from African ancestry cohorts (AWI-Gen and TrypanoGEN+) by geographical region of the African continent.** N.A. indicates samples missing country of origin information.
